# Evaluation of County-Level Heterogeneity in Excess Mortality in Colorado from March to September 2020

**DOI:** 10.1101/2021.04.10.21255235

**Authors:** Jay Chandra, Marie Charpignon, Mathew C. Samuel, Anushka Bhaskar, Saketh Sundar, Kirk Bol, Yuan Lai, Leo A. Celi, Sema K. Sgaier, Grace Charles, Maimuna S. Majumder

## Abstract

1.

**Importance:** Tracking the direct and indirect impact of the coronavirus disease 2019 (COVID-19) pandemic on all-cause mortality in the United States has been hindered by the lack of testing and by reporting delays. Evaluating excess mortality, or the number of deaths above what is expected in a given time period, provides critical insights into the true burden of the COVID-19 pandemic caused by the novel Severe Acute Respiratory Syndrome Coronavirus 2 (SARS-CoV-2). Stratifying mortality data by demographics such as age, sex, race, ethnicity, and geography helps quantify how subgroups of the population have been differentially affected. Similarly, stratifying mortality data by cause of death reveals the public health effects of the pandemic in terms of other acute and chronic diseases.

**Objective:** To provide stratified estimates of excess mortality in Colorado from March to September 2020.

**Design, Setting, and Population:** This study evaluated the number of excess deaths both directly due to SARS-CoV-2 infection and from all other causes between March and September 2020 at the county level in Colorado. Data were obtained from the Vital Statistics Program at the Colorado Department of Public Health and Environment. These estimates of excess mortality were derived by comparing population-adjusted mortality rates in 2020 with rates in the same months from 2015 to 2019.

**Results:** We found evidence of excess mortality in Colorado between March and September 2020. Two peaks in excess deaths from all causes were recorded in the state, one mid-April and the other at the end of June. Since the first documented SARS-CoV-2 infection on March 5th, we estimated that the excess mortality rate in Colorado was two times higher than the officially reported COVID-19 mortality rate. State-level cumulative excess mortality from all causes reached 71 excess deaths per 100k residents (∼4000 excess deaths in the state); in contrast, 35 deaths per 100k directly due to SARS-CoV-2 were recorded in the same period (∼1980 deaths. Excess mortality occurred in 52 of 64 counties, accounting for 99% of the state’s population. Most excess deaths recorded from March to September 2020 were associated with acute events (estimated at 44 excess deaths per 100k residents and at 9 after excluding deaths directly due to SARS-CoV-2) rather than with chronic conditions (∼21 excess deaths per 100k). Among Coloradans aged 14-44, 1.4 times more deaths occurred in those months than during the same period in the five previous years. Hispanic White males died of COVID-19 at the highest rate during this time (∼90 deaths from COVID-19 per 100k residents); however, Non-Hispanic Black/African American males were the most affected in terms of overall excess mortality (∼204 excess deaths per 100k). Beyond inequalities in COVID-19 mortality per se, these findings signal considerable regional and racial-ethnic disparities in excess all-cause mortality that need to be addressed for a just recovery and in future public health crises.

## 2. Introduction

Since December 2019, the rapid unfolding of the coronavirus disease 2019 (COVID-19) pandemic, coupled with a lack of preparedness across the country has led to complications in understanding its true impact. In the initial phases of the pandemic, the number of laboratory-confirmed cases could only account for 10-15% of all the severe acute respiratory syndrome coronavirus 2 (SARS-CoV-2) infections in existence [1]. In contrast, deaths attributable to a SARS-CoV-2 infection were much more likely to be accounted for and documented. Because of the burden COVID-19 has placed on healthcare systems, there remains a need to quantify deaths from other causes that occurred during the pandemic to evaluate the overall mortality effects of the novel infection.

To motivate our examination of all-cause mortality during the pandemic, we defined four categories of deaths: direct counted, indirect counted, directed uncounted, and indirect uncounted deaths [2]. *Direct counted deaths* are those where COVID-19 is listed on the death certificate as the underlying cause of death and is determined by the presence of a positive test. This metric can be challenging due to limitations in testing capacity and missed diagnoses earlier in the pandemic. For *indirect counted deaths*, a COVID-19 test was administered and yielded a positive result, but the infection was listed on the certificate as a contributing (secondary) – but not as an underlying (primary) – cause of death (e.g., patient with end-stage metastatic breast cancer who got infected with SARS-CoV-2 by a visitor while on therapy at the hospital and subsequently died from respiratory and immune system complications). *Direct uncounted deaths* correspond to situations where COVID-19 was not listed on the death certificate (i.e., neither as an underlying nor contributing cause of death) *though the infection was probable*. This scenario, while rare, might have occurred in the early stages of the pandemic because of inadequate testing and a lack of specific guidelines geared towards medical certifiers in the absence of confirmatory laboratory results. For *indirect uncounted deaths*, no COVID-19 test was administered, and the infection was not suspected to be responsible for the fatal event. Therefore, COVID-19 was not listed on the death certificate as an underlying or contributing cause of death. An example of such a scenario would be a patient hospitalized with a stroke who did not receive adequate care because the hospital was overwhelmed by COVID-19 cases and died at the institution during their stay.

Two situations complicate the estimation of mortality *directly* and *indirectly* due to SARS-CoV-2 infection. First, if an individual died of COVID-19 but did not receive a positive test result (due to insufficient testing or inaccuracy of the test itself), their death may not be counted towards *direct* COVID-19 deaths (i.e., a false negative). Second, if an individual died from another underlying cause but tested positive for COVID-19 during a routine screening, then their death *would be* counted towards *indirect* COVID-19 deaths in national health statistics (i.e., a false positive). Therefore, relying solely on *direct* or *counted* COVID-19 deaths may be an unreliable indicator of overall COVID-19-related mortality, which should also include *indirect* and *uncounted* COVID-19 deaths.

While *direct* and *indirect counted* COVID-19 deaths have been used to quickly assess mortality directly due to SARS-CoV-2 infection, this approach has several limitations. Data used to calculate case fatality rates (i.e., the percentage of documented SARS-CoV-2 infections resulting in death) are generally based on a first report of a death – by healthcare providers, facilities, families, or through routine contact tracing. Thus, official causes of death may not be known immediately. In contrast, direct linkage to state-level death certificate-based data [3, 4] collected through vital records registration, or to the National Death Index hosted by the National Center for Health Statistics, would make such cause-of-death information available. These data linkages also provide access to the detailed list of factors that significantly contributed to the death, beyond the underlying cause.

In the recent past, excess mortality calculations have been used to better encapsulate deaths attributable to natural disasters [5], wars, and infectious disease outbreaks, including national and state-level estimates for COVID-19 [6]. This approach is adapted to quantify the *direct* and *indirect* impacts of the disease on both all-cause and cause-specific rates [7], i.e., either on mortality in aggregate or by disease (e.g., cancer, diabetes). Furthermore, geographically stratifying mortality data can help public health officials and policymakers identify demographics most impacted by the COVID-19 pandemic and mobilize resource allocation accordingly. In particular, county-level analysis can yield insights into heterogeneous impacts on excess mortality within a population of interest. Counties are also a relevant geographical unit of analysis because local-level public health agencies, prevention activities, and interventions operate at this level [8].

In March 2020, we began working with the Colorado Department of Public Health and Environment (CDPHE) to estimate the true burden of COVID-19 on all-cause mortality throughout the state at the county level. Testing in Colorado was limited in the early months of the pandemic, and the state experienced a number of outbreaks in nursing homes, restaurants, prisons, and meatpacking plants [9]. We obtained detailed mortality data for 2015 to 2020 from the CDPHE. Data were stratified by age, race, sex, geography, and cause of death, allowing us to quantify the toll of the pandemic on all-cause mortality for different population subgroups.

## 3. Methods

### 3.1. Data

The Vital Statistics Program at the CDPHE provided our team with de-identified death certificate data from January 1st of 2015 to September 10th of 2020. These individual-level records included the ICD-10 code(s) defining the underlying cause(s) of death, additional ICD-10 code(s) for contributing cause(s) of death, facility in which the patient died, race (White, Black/African American, Asian/Pacific Islander, and American Indian/Alaska Native), ethnicity (i.e., Hispanic and Non-Hispanic groups), age group, sex, county of death, county of residence, and week of death in the year. The data included both underlying (i.e., primary) and contributing (i.e., secondary) cause of death, grouped into the following categories: COVID-19, motor vehicle accident, work-related accident, other accident, drug overdose, suicide, homicide, influenza, pneumonia, cancer, heart disease, cerebrovascular disease, chronic obstructive pulmonary disease (COPD), diabetes, liver disease, Alzheimer’s disease, and other (see Supp Table 2 for the complete list of ICD-10 codes). To obtain population-normalized all-cause and cause-specific mortality estimates, we used population data from the Colorado State demography office stratified by county, sex, age, race, and ethnicity, for years ranging from 2014 to 2019 [10]. See Supp Materials 8 for details on the timeliness of death certificate data.

While open access datasets available to the general public, such as the repository operated by Johns Hopkins University Center for Systems Science and Engineering (JHU CSSE [3]), provide insight into direct and indirect counted deaths, death certificates are a more reliable source of information due to the detailed assessment of the medical examiner. Such data enables researchers to better understand subtleties of what is regarded as a COVID-19 death from both underlying and contributing causes of death [4]. As an example of this nuance, studies released so far have highlighted a high prevalence of cardiovascular disease in patients with COVID-19, but many patients also develop heart injury from COVID-19 [11]. In both cases, COVID-19 and heart disease could be either the underlying or a contributing cause of death. To contrast estimates of deaths directly due to SARS-CoV-2 infection as certified by the CDPHE against open access datasets available to the general public, we used the COVID-19 death time series from the JHU CSSE (Supp Materials 1). We conducted a longitudinal comparison of the two data sources at both the state (Section 4.1) and county level (Supp Materials 2, Supp Fig. 1, Supp Fig. 2).

### 3.2. Definition of excess mortality

The most common approach to the calculation of excess mortality compares the number of deaths from all causes in a given period to expected deaths, as determined using historical data [12]. Here, we defined excess mortality as the difference between deaths that occurred in 2020 and the average of deaths reported from 2015 to 2019, consistent with the termed empirical baseline described by Weinberger et al. in their state-level excess mortality work [4] (Supp Materials 3). It is important to note, however, that solely considering the magnitude of excess deaths could obscure the nuances of COVID-19 impact on all-cause mortality among underrepresented or small population subgroups. In such cases, studying the ratio of deaths in 2020 as compared to the empirical baseline from 2015 to 2019 may provide further clarity and allow for additional comparisons between age groups and sex, race, and ethnicity subgroups. In analyses stratified by sex, race, and ethnicity, we computed age-adjusted death rates [13, 14], using the 2000 US Standard Population weights published by the CDC [15, 16], to account for any potential difference in age distributions and provide comparable estimates across population subgroups. Confidence intervals (CI) for excess mortality and ratios were defined using a t-distribution with empirical death data from 2015 to 2019 (Supp Materials 4).

### 3.3. County- and CBSA-level excess mortality analysis

To better understand the heterogeneous impact of COVID-19 across the state of Colorado, we examined excess mortality at both the county and Core Based Statistical Area (CBSA) level. Further, our analysis incorporated an assessment of deaths directly due to SARS-CoV-2 infection and excess all-cause mortality by county and CBSA typology. Each county is classified according to the Colorado Rural Health Center as urban, rural, or frontier [17]. An urban county is defined as having a high degree of economic and social integration with a metropolitan area. Rural counties are those that are not linked to any metropolitan area. Finally, frontier counties are highly isolated from population centers and services (six or fewer persons per square mile). Similarly, CBSAs can be of two types: metropolitan or micropolitan. For details about the benefits of CBSA-level analyses to inform public health policies and the analyses themselves, refer to Supp. Materials 5 and Supp Fig. 3 for CBSA excess all-cause mortality results. In addition to a cross-sectional analysis, we conducted a longitudinal analysis to account for differences in the timing of the pandemic across geographies. For details about monthly excess deaths, refer to Supp. Materials 5 and Supp Figs. 4-5.

### 3.4. Places of death and causes of death

Using death certificates, we quantified the change in deaths occurring in each type of facility for the state of Colorado from 2015 to 2020: home residence, hospital/inpatient, nursing home, outpatient/emergency room (ER), hospice, and other (which may include workplace, another person’s residence, outdoor location, commercial building, etc.). Then, starting from the week of March 5th until the week ending on August 27th, we quantified the change in cause-specific deaths in Colorado from 2015 to 2020. We grouped the aforementioned ICD-10 codes into two categories: acute and chronic causes of death. The acute category encompasses the following fatal events: motor vehicle accident, work-related accident, other accident, drug overdose, suicide, homicide, influenza, and pneumonia. The chronic category includes cancer, heart disease, cerebrovascular disease, COPD, diabetes, liver disease, and Alzheimer’s disease. This set of ICD-10 codes includes many of the top causes of death in Colorado [18] and accounts for 78% of the total deaths in 2020 (Supp Table 2).

### 3.5. Sensitivity analyses

#### 3.5.1. Adjustments for the timing of community transmission

In addition to the main analysis, we conducted a sensitivity analysis with both an early and a late starting point to estimate excess deaths during the pandemic in Colorado, at the state, county, and CBSA level. The main analysis was conducted using death counts as estimated since the first documented infection in Colorado (March 5th). We conducted two sensitivity analyses against this start date, using the date of the first case in the state +/−28 days. Considering an earlier date (February 6th) as a sensitivity analysis enabled us to investigate the possibility that COVID-19 was affecting Coloradoans before the record of the first documented infection. To test the consistency and robustness of our estimates, we also evaluated the effect of choosing a later date (April 2nd), corresponding to the period when the cumulative number of deaths directly attributed to SARS-CoV-2 infection exceeded the expected number of deaths in a week in Colorado. Results from these analyses are presented in Supp Fig. 6.

#### 3.5.2. Adjustments for time-varying population size

Excess deaths were calculated for each geographical level (i.e., county, CBSA, and state) in Colorado and normalized by population size. Two different methods were used to account for the size of the population per county. The first approach used county and CBSA population sizes that were constant over time. After calculating excess deaths as described above, we divided these quantities by the population of each geographical area as estimated in 2017. Given that the last national census was conducted in 2010, population sizes used in this study are projected estimates rather than empirical ones. Results presented in Section 4 rely on this approach. To quantify the effect of a time-varying county population size on excess all-cause mortality estimation at these levels and thereby assess the robustness of our results, a variable population size approach is presented as a sensitivity analysis (Supp Materials 6, Supp Fig. 7). In both approaches, population-normalized death counts from years 2015 to 2019 were averaged and subtracted from 2020 population-normalized deaths, yielding population-normalized excess deaths.

## 4. Results

### 4.1. Peaks of excess all-cause mortality during the pandemic

Between March and September 2020, there was evidence of excess deaths in Colorado. Two peaks were recorded at the state level, one around mid-April and the other towards the end of June. Since the first documented infection on March 5th, we estimate that state-level cumulative excess mortality across all causes reached a magnitude of 71 excess deaths per 100k residents. This translates to approximately 4000 excess deaths in Colorado from March to September 2020, i.e., 21% of total deaths during the period. As a comparison, there were 1980 deaths directly due to SARS-CoV-2 infection during the same period, i.e., 10.4% of total deaths. For details about the underlying structure of the Colorado population, refer to Supp Materials 7 and Supp Table 1.

A descriptive longitudinal analysis of state-level data revealed the peaks and temporal variation of excess all-cause mortality in Colorado. Overall, we estimated 71 excess deaths per 100k residents (∼4000 excess deaths at the state level). We observed a peak in excess deaths from April 11th to 17th (5.8 excess deaths per 100k residents), one week before the peak in deaths directly due to SARS-CoV-2 infection. From April 11th onwards, there were 63 excess deaths per 100k residents (∼3500 excess deaths at the state level). However, using February 6th instead of March 5th as the start date of the analysis yielded 76 excess deaths per 100k residents (∼4300 excess deaths at the state level) over the considered time period.

In May 2020, the overall mortality burden caused by COVID-19 was still substantial, with 11.4 excess deaths per 100k residents registered in the state (11.2 deaths per 100k directly due to SARS-CoV-2 infection). A second peak in weekly excess all-cause mortality was detected from June 20th to 26th (3.4 excess deaths per 100k residents), though its magnitude was lower than that of the first peak (Fig. 1a). In August, deaths directly due to SARS-CoV-2 infection decreased to levels seen in March, with 1.8 deaths per 100k residents at the state level. However, excess all-cause mortality remained high, with 10.2 excess deaths per 100k residents in comparing 2020 to the baseline period of 2015 to 2019.

As time progressed, the difference between cumulative excess deaths from all causes and deaths directly due to SARS-CoV-2 infection accelerated. Initially estimated at 15.5 deaths per 100k residents (April 11th-17th), the difference rose to 31 deaths per 100k (June 20th-26th) and then reached 48 deaths per 100k (August 29th-September 4th) (Fig. 1b). For context, 135, 275, and 396 deaths from all causes were recorded over the same time intervals from 2015 to 2019 on average.

**Figure 1.**
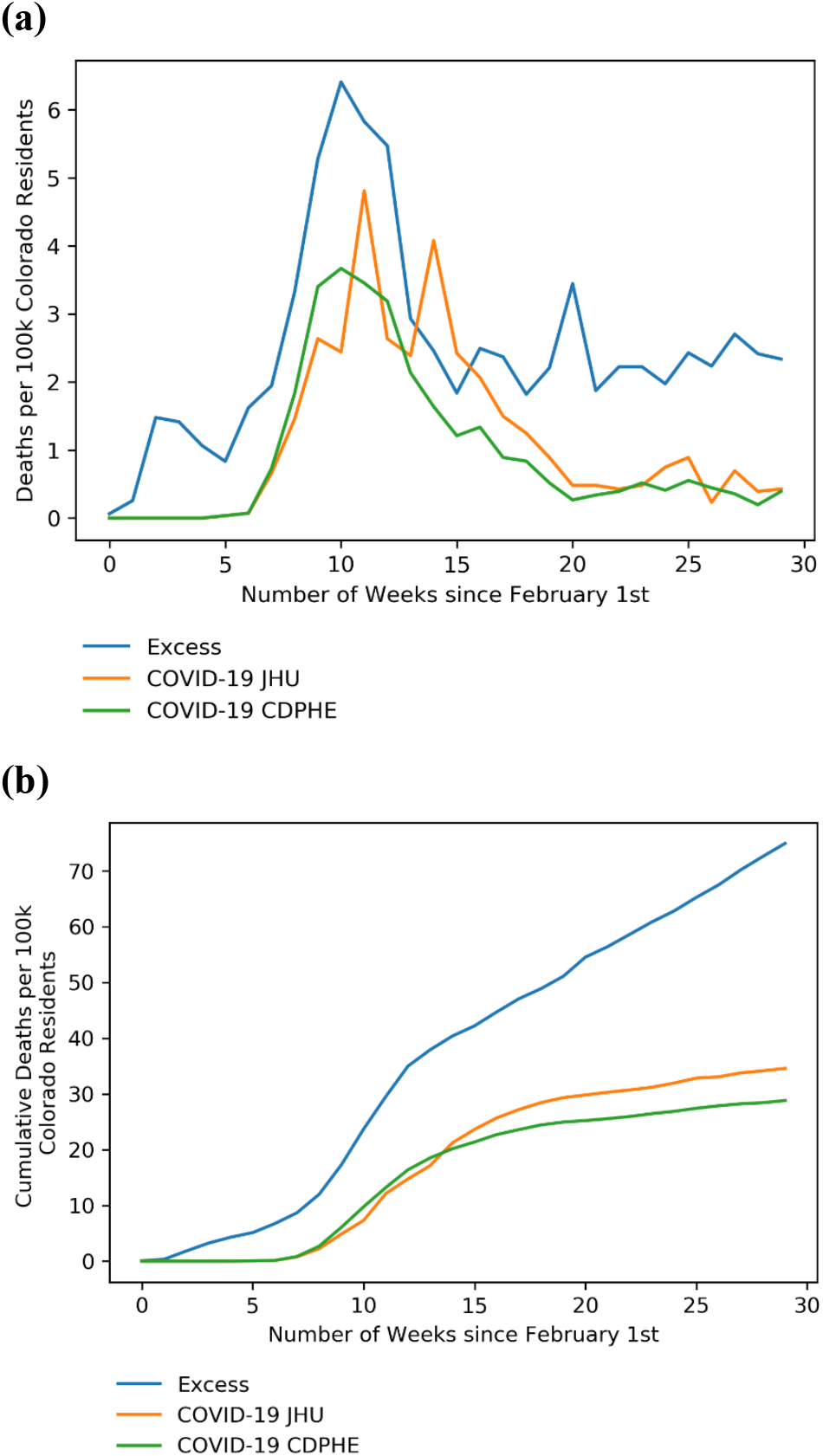
Weekly excess deaths and COVID-19 deaths among residents, from February 1st to September 6th. **(a)** Weekly deaths recorded since February 1st, for each of the three time series. **(b)** Cumulative deaths recorded since February 1st, for each of the three time series. For both **(a)** and **(b)**, the blue line represents the estimated excess deaths among residents, normalized to the state’s population size projection in 2017. The orange line represents the COVID-19 deaths in Colorado as reported by the JHU COVID-19 data repository, normalized similarly. The green line represents the deaths directly due to SARS-CoV-2 infection in the state as reported by the Colorado CDPHE, normalized similarly.

### 4.2. Excess all-cause mortality by county and over time

From March 5th to September 10th, 52 out of 64 counties (i.e., 81% of counties, representing 99% of the Colorado population) presented excess all-cause mortality, including 94% of urban, 83% of rural, and 69% of frontier counties.

#### 4.2.1. Geographical differences in excess all-cause mortality

A three-part scatter plot represents population-normalized COVID-19 (as determined by death certificates provided from the CDPHE) and excess deaths in 2020, as compared with the 2015 to 2019 average, rendering disparities across county types. Comparing the subset of deaths directly due to SARS-CoV-2 infection to deaths from all causes sheds light on the downstream consequences of the COVID-19 pandemic on overall mortality trends. Based on the level of excess all-cause mortality relative to COVID-19 mortality alone, three clusters – labeled A, B, and C – were delimited in Figures 1 to 4. Cluster C includes counties that recorded fewer deaths in 2020 than in the five previous years, while clusters A and B include counties that presented more deaths in 2020 than in years prior. Specifically, the number of excess all-cause deaths was higher than the number of deaths directly due to SARS-CoV-2 infection in 72% of counties (cluster A). In contrast, only 9% of counties showed the opposite pattern, i.e., deaths directly due to SARS-CoV-2 infection outnumbered excess deaths (cluster B). Finally, 19% of counties had fewer reported deaths than during the five previous years (cluster C), though this must be interpreted with caution, as most counties belonging to this cluster have small population sizes (median of 5900 residents) (Fig. 2).

**Figure 2.**
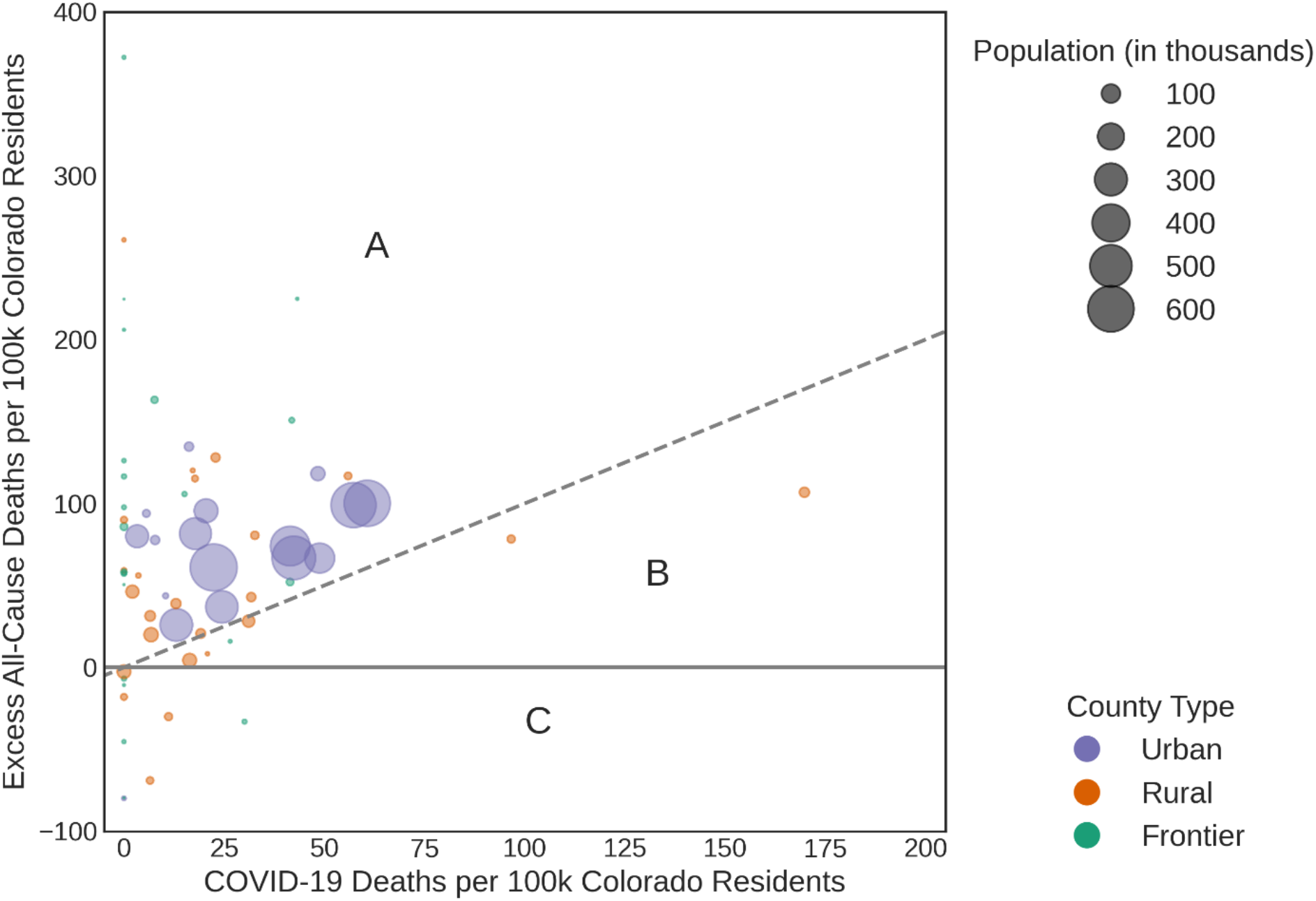
Overall county-level excess all-cause mortality vs. deaths directly due to SARS-CoV-2 infection among residents. Region A: More deaths in 2020 than in prior years, excess deaths > COVID-19 deaths. Region B: More deaths in 2020 than in prior years, COVID-19 deaths > excess deaths. Region C: Less deaths in 2020 than in prior years. Counts correspond to the period starting on the date of the first documented infection in the state (March 5th) and ending on September 10th. Population-normalized estimates were computed using county-level population projections from 2017. Cluster A represents the counties of Colorado that had more population-normalized excess deaths from all causes in 2020 than fatal events primarily attributed to COVID-19 by medical certifiers. Cluster B is composed of counties where the COVID-19 death rate exceeded the value of population-normalized excess all-cause mortality rate. Counties falling into cluster C were characterized by negative excess deaths in 2020. The points were sized in proportion to the population of each county and were colored based on the county type (i.e., urban in purple, rural in orange, and frontier in green).

#### 4.2.2. Temporal evolution of excess mortality

Over time, the difference between cumulative deaths directly due to SARS-CoV-2 infection and excess deaths was mainly observed in urban counties, as compared to rural and frontier areas of Colorado, where the difference remained constant over time. In March 2020, 39 out of 64 counties experienced excess mortality (i.e., 61% of counties, representing 95% of the Colorado population), including 94% in urban, 58% in rural areas, and 39% in frontier areas. During that month, we estimated only about 1.2 deaths per 100k residents directly due to SARS-CoV-2 infection and 6.4 excess all-cause deaths per 100k. Yet, by April, there was increasing evidence of the disruption caused by SARS-CoV-2 transmission across all counties of Colorado, regardless of their typology. At the state level, 12.4 deaths per 100k residents directly due to SARS-CoV-2 infection were recorded; in the most impacted area, Chaffee County, this rate reached up to 72 deaths per 100k. Excess all-cause mortality was the highest in April, reaching 24 excess deaths per 100k residents at the state-level and peaking at 107 excess deaths per 100k in Dolores County. In June, both deaths directly attributed to SARS-CoV-2 infection (4.1 deaths per 100k) and excess all-cause mortality (8.5 excess deaths per 100k) decreased significantly as compared to previous pandemic months. In July, however, while deaths directly due to SARS-CoV-2 infection plateaued at 5.0 per 100k residents, excess deaths continued to increase in the state at a rate of 11.5 per 100k (Supp Fig. 3). Overall, except for three outliers (one urban: Broomfield; two rural: Chaffee and Otero), excess deaths recorded from March to September 2020 followed the same trend as deaths directly due to SARS-CoV-2 infection (Spearman rank correlation: 0.62, Pearson rank correlation: 0.67).

### 4.3. Disparities in excess all-cause mortality by sociodemographics

#### 4.3.1. Excess all-cause mortality by age

Across the state of Colorado, adults aged 85+ had the highest rate of excess all-cause mortality from March 5th to September 10th (1070 excess deaths per 100k [95% CI: 984;1150]), followed by those aged 75-84 (460 excess deaths per 100k [95% CI: 360;556]) and 65-74 (200 excess deaths per 100k [95% CI: 144;256]). The 1-4 age group had negative excess deaths at the state level (Supp Fig. 6a). In addition, the age-stratified ratio of all-cause deaths recorded in 2020 to the average during the same period in 2015 to 2019 revealed that more fatal events than observed in the five previous years were recorded among Coloradans aged 15-24 (1.36 times more [95% CI:1.28;1.44]), 25-34 (1.39 times more [95% CI: 1.28;1.50]), and 35-44 (1.39 times more [95% CI: 1.24;1.54]), followed by the 65-74 age group (1.28 times more [95% CI: 1.18;1.38]) (Supp Fig. 6b).

#### 4.3.2. COVID-19 and excess all-cause mortality by sex, race, and ethnicity

We found that Hispanic White males died of COVID-19 at the highest rate (90 deaths per 100k directly due to SARS-CoV-2 infection), while Non-Hispanic Black/African American (BAA) males were also severely affected (85 deaths per 100k), followed by Non-Hispanic Asian/Pacific Islander males (60 deaths per 100k). In contrast, the COVID-19 mortality rate among Non-Hispanic White males was estimated at 25 deaths per 100k, the lowest rate among males (Supp Fig. 8). Similarly, we found that Non-Hispanic BAA males experienced higher rates of excess all-cause mortality, with 201 excess deaths per 100k [95% CI: 145;257] as compared to the 2015 to 2019 average (i.e., 1.44 times the baseline mortality rate [95% CI: 1.28;1.60]). In the same racial-ethnic group, females were less severely impacted, with an excess all-cause mortality rate of 152 deaths per 100k [95% CI: 137;167]. Among males, Hispanic White and Non-Hispanic Asian/Pacific Islander communities were anomalously vulnerable to excess all-cause mortality, with rates reaching 185 [95% CI: 168;202] and 152 [95% CI: 126;178] excess deaths per 100k residents (Fig. 3a). The age-adjusted ratio of death counts in 2020 vs. 2015 to 2019 ranged from 1.09 to 3.2, highlighting significant health disparities between subgroups of the population, as defined by their sex, race, and ethnicity. Irrespective of sex and race, those who identified as ethnically Hispanic were the most affected (Hispanic BAA females: 3.24 times more deaths in 2020 vs. 2015 to 2019 [95% CI: 2.27;4.21], Hispanic Asian/Pacific Islander males: 3.44 [95% CI: 1.47;5.41]). Notably, although females overall were less impacted in terms of excess mortality, the subgroup of Non-White Hispanic females experienced an all-cause mortality rate 1.63 [95% CI: 1.21;2.05] times higher than observed in the last five years (Fig. 3b).

**Figure 3.**
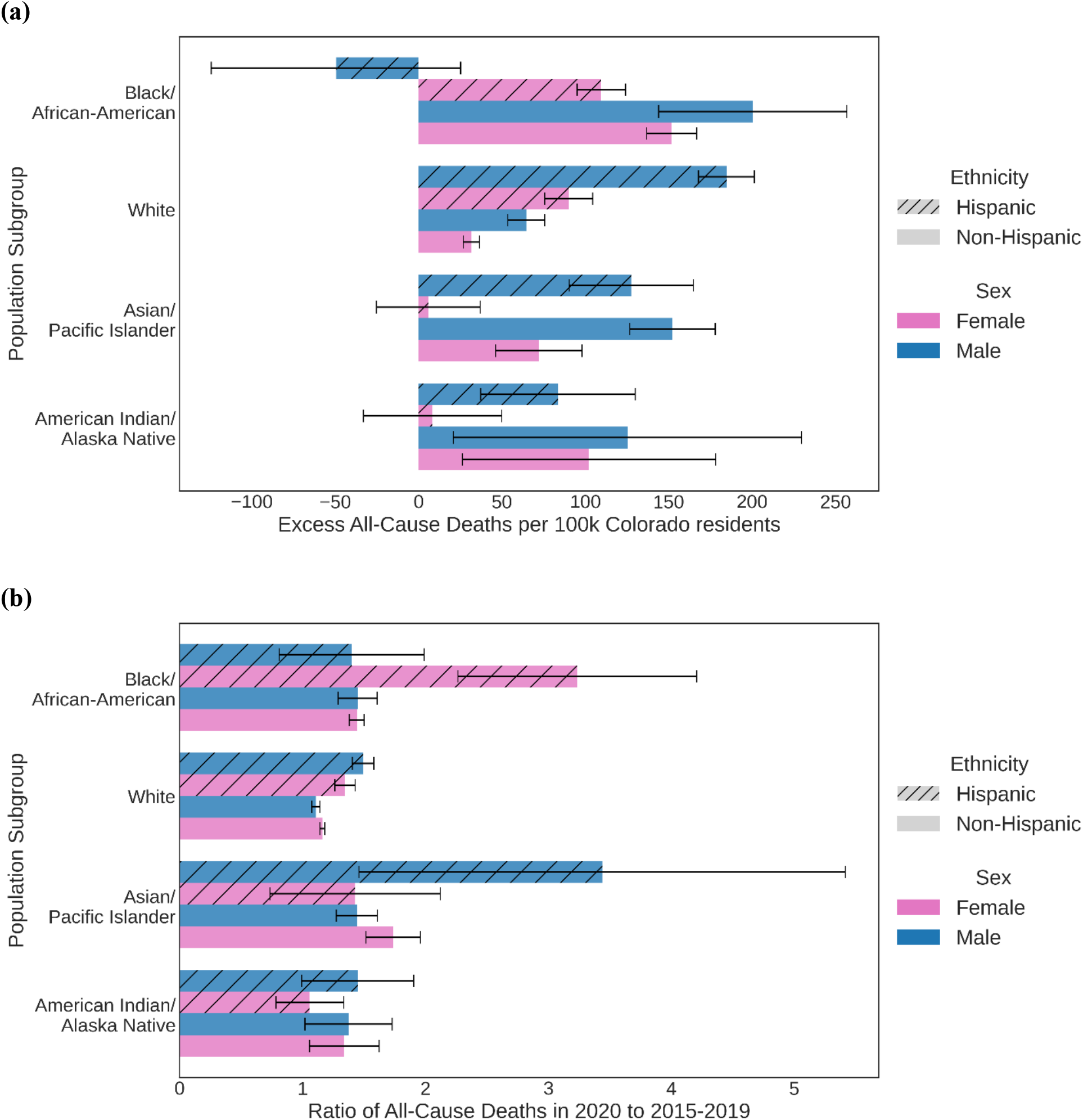
Age-adjusted excess all-cause mortality among residents, stratified by (race, ethnicity, sex) population subgroup. Counts correspond to the period starting on the date of the first documented infection in the state (March 5th) and ending on September 10th. Population-normalized estimates were computed using state-level population projections from 2017. **(a)** Age-adjusted and population-normalized excess deaths and **(b)** ratio of all-cause deaths in 2020 to the average all-cause deaths from 2015 to 2019 are shown for each (race, ethnicity, sex) population subgroup in Colorado. Hashed and solid bars represent Hispanic ethnicity, while solid bars represent Non-Hispanic ethnicity. Blue and pink bars represent male and female, respectively. Vertically, results are grouped by race (Black/African American, White, Asian/Pacific Islander, and American Indian/Alaska Native).

#### 4.3.3. Excess all-cause mortality by cause of death

Beyond age, sex, race, and ethnicity, examining mortality by underlying cause of death can yield additional insights. From the week of March 5th to the week ending on August 27th, we observed maximal excess deaths from heart disease (8.4 excess deaths per 100k [95% CI: 5.5;11.3]) and drug overdose (5.4 excess deaths per 100k [95% CI: 4.8;6.0]) as the underlying cause of death. Additionally, the number of deaths attributed to Alzheimer’s disease as an underlying cause increased from March to August 2020 as compared to the same period in 2015 to 2019 (4.1 excess deaths per 100k [95% CI: 2.8;5.5]). Conversely, there were fewer deaths directly due to pulmonary diseases (i.e., influenza, pneumonia, and COPD) in 2020 than in the five previous years (−0.16 [95% CI: −0.46;0.14], −0.47 [95% CI: −1.39;0.45], and −1.1 [95% CI: −1.7;-0.6] excess deaths per 100k, respectively). By classifying the underlying causes of death into two categories (acute and chronic), we estimated 20.5 [95% CI: 14.5;26.5] and 9.0 [95% CI: 6.6;11.4] excess deaths per 100k for acute and chronic conditions, respectively (Fig. 4a). In addition, the ratio of cause-specific mortality from March 5th to September 10th revealed that significantly more deaths than observed in the five previous years were recorded for drug overdose (1.76 times more [95% CI:1.60;1.92]), homicide (1.54 times more [95% CI: 1.21;1.88]), and 15 other causes of death (6 from acute and 9 from chronic medical conditions). Consistent with our previous analysis, causes of death related to pulmonary health all had ratios less than one (Fig. 4b).

**Figure 4.**
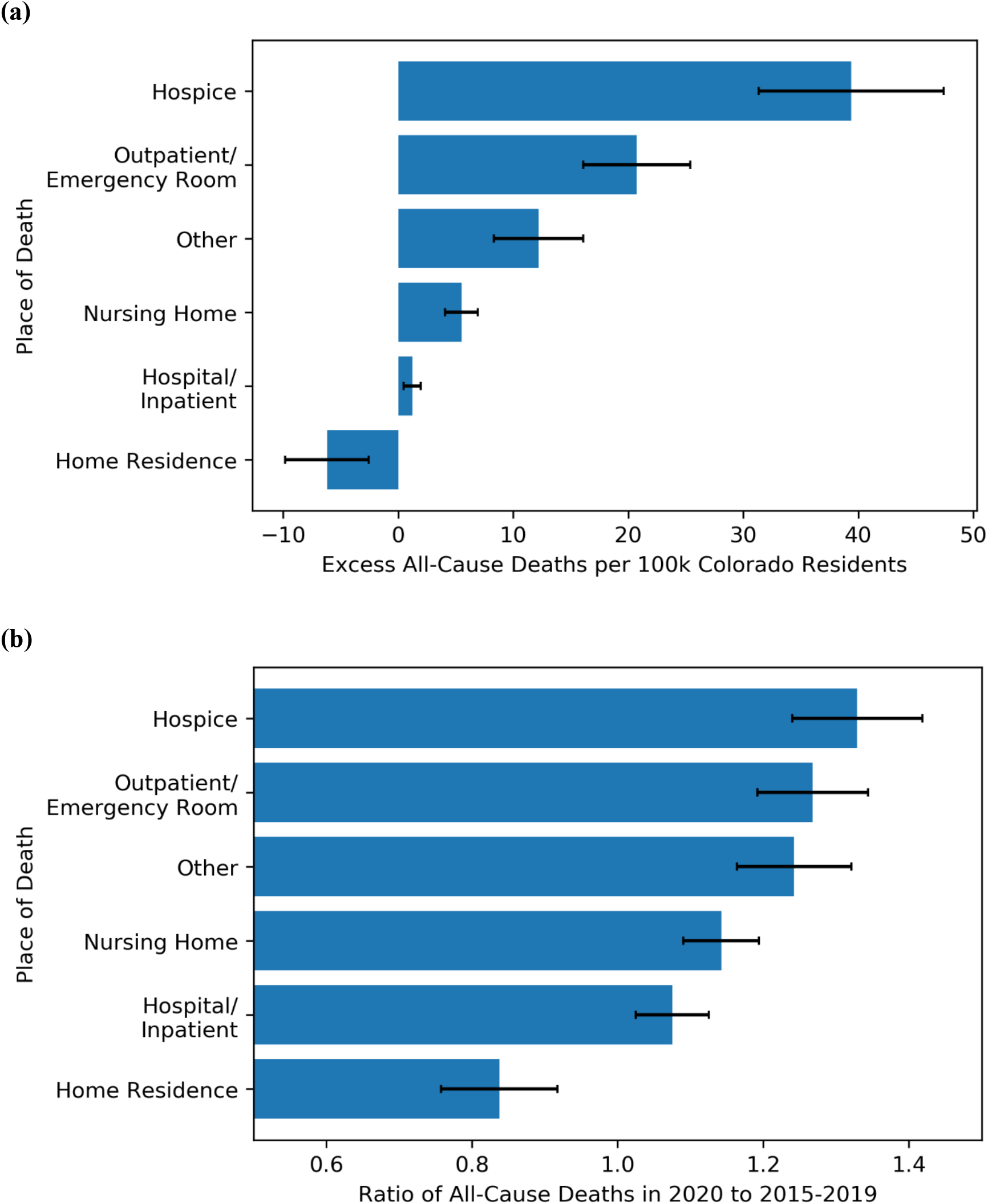
Deaths among residents, stratified by place of death. Counts correspond to the period starting on the date of the first documented infection in the state (March 5th) and ending on September 10th. Places of death are separated into six categories as denoted on the death certificates: home residence, hospital/inpatient, nursing home, outpatient/emergency room (ER), hospice, and other (which may include workplace, another person’s residence, outdoor location, commercial building, etc.). The results are ordered by **(a)** the excess mortality estimated value, from the highest at the top (Home Residence: 37.6 excess deaths per 100k) to the lowest at the bottom (Hospice: - 5.8 excess deaths per 100k) and **(b)** the ratio of 2020 all-cause deaths to the 2015 to 2019 average deaths, from the highest at the top (Home Residence: 1.3) to the lowest at the bottom (Hospice: 0.8).

#### 4.3.4. Excess all-cause mortality by place of death

System-based factors also influence the likelihood of home deaths [19]. Dying in a preferred place may be a quality marker for end-of-life care among older populations [20]. However, it can also signal a fragile health system unequipped to resist repeated surges and sometimes avoided – leaving younger people undiagnosed and possibly dying at home. From March to September 2020, deaths at home alone accounted for 52% of excess deaths recorded. Our analysis of all-cause mortality in Colorado showed that excess deaths were the highest at home (39.4 per 100k [95% CI: 31.4;47.5]), followed by the hospital (20.7 per 100k [95% CI: 16.7;24.8]), and nursing homes (12.2 per 100k [95% CI: 8.3;16.1]) (Fig. 5a). This implies that there were 1.3 times more deaths from all causes that occurred at home than in the five previous years during the same period (Fig. 5b). While nursing homes were the third highest setting associated with an excess all-cause mortality rate in Colorado, hospices had less deaths in 2020 than in 2015 to 2019.

**Figure 5.**
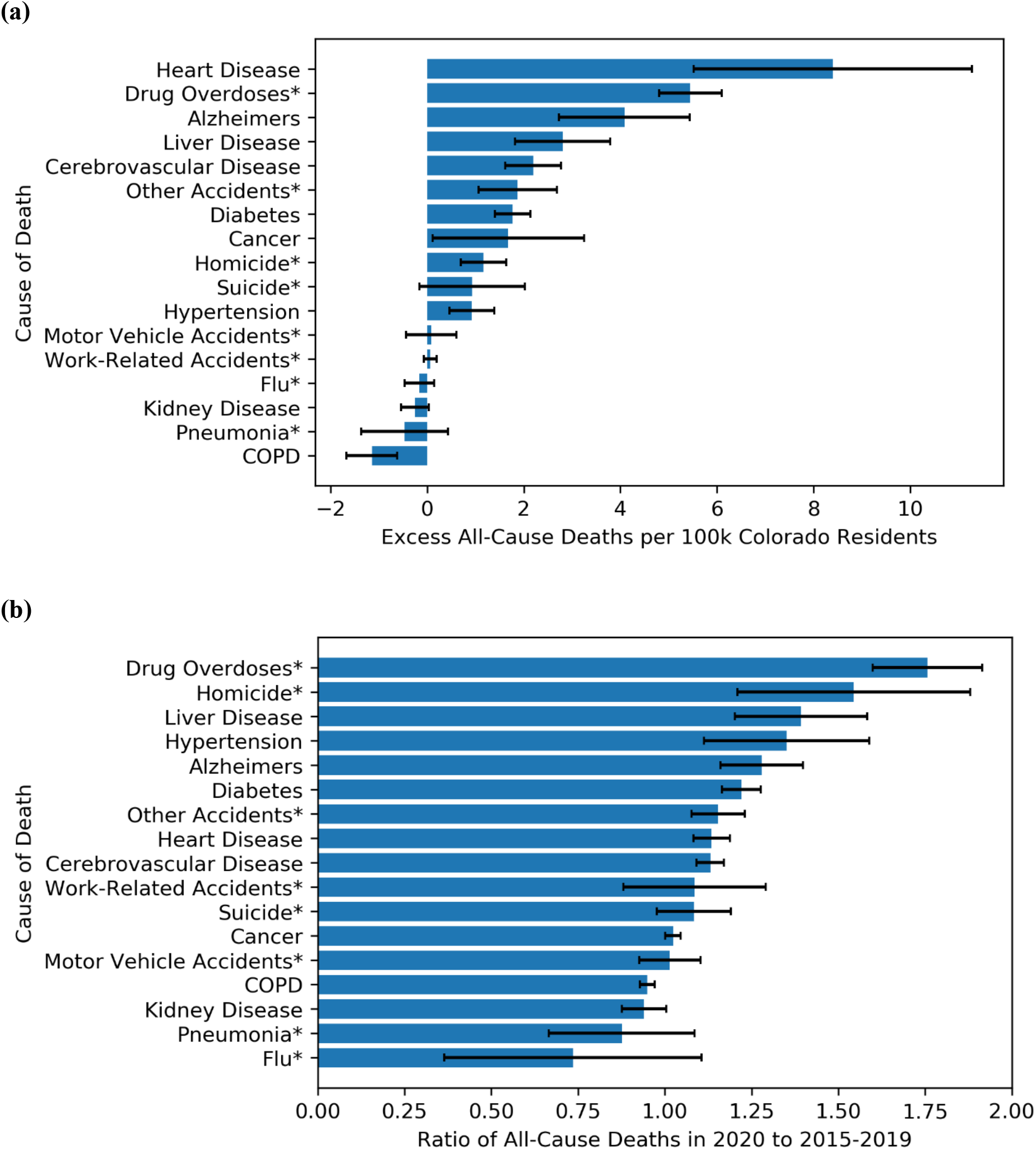
Deaths among residents, stratified by underlying cause. Counts correspond to the period starting on the date of the first documented infection in the state (March 5th) and ending on September 10th. Population-normalized estimates were computed using state-level population projections from 2017. Excess death estimates for 17 major underlying causes of death in Colorado are presented. The cause-specific results (as defined by the underlying cause indicated on the death certificate) are ordered by **(a)** the excess mortality estimated value, from the highest at the top (heart disease: 8.4 excess deaths per 100k) to the lowest at the bottom (COPD: −1.2 excess deaths per 100k) and **(b)** the ratio of cause-specific deaths in 2020 to the average cause-specific deaths from 2015 to 2019, from the highest at the top (drug overdoses: 1.76) to the lowest at the bottom (Flu: 0.74). The presence of an asterisk denotes acute deaths, while its absence refers to chronic deaths.

## 5. Discussion

Our findings indicate that across both acute and chronic medical conditions, there has been a vast undercounting of direct and indirect COVID-19-related deaths in Colorado. Excess deaths increased by 59% from March to September 2020 (from 6.4 to 10.2 per 100k residents), peaking at 275% in April 2020 (24.0 per 100k residents). The peak of excess deaths was observed one week before the peak in deaths directly due to SARS-CoV-2 infection (April 18th-24th of 2020), suggesting that potential COVID-19 deaths were neither diagnosed nor reported early on in the pandemic. Reasons for this discrepancy could be either that the lack of testing capacity led to reporting inaccuracies of mortality cause(s) on death certificates, or that the medical examiner identified COVID-19 as a contributing cause, but not as the underlying (i.e., direct) cause of death. Given the lack of institutional recommendations regarding the use of ICD-10 codes for SARS-CoV-2 infection prior to their release by the WHO on April 20th [21], certifying authorities may have chosen to instead report other underlying causes of death (e.g., influenza or pneumonia) for fatal events that occurred in March and April. Another explanation could be that Coloradans were dying for other reasons such as health insurance disruption or reluctance to seek care associated with fear of infection with SARS-CoV-2. Additionally, shelter-at-home policies might have led to increased instances of substance abuse, suicide, or domestic violence. Among all causes of death, drug overdoses, liver disease, and Alzheimer’s disease registered the largest degree of excess mortality. Conversely, there were fewer deaths from influenza, pneumonia, and COPD than in the previous five years. This may be explained by the effectiveness of non-pharmaceutical interventions (NPI) at preventing infectious respiratory diseases other than SARS-CoV-2.

Beyond understanding the temporality of excess all-cause mortality, we were particularly interested in distinguishing fatal events associated with outbreaks (e.g., in nursing homes) from other acute (e.g., death from hemorrhage in the ER) or chronic fatal events (e.g., death at home attributed to neurological disease) that the pandemic might have precipitated. To that end, we examined the distribution of excess all-cause mortality by place of death, shedding light on how Colorado’s healthcare infrastructure has been utilized during the pandemic. Importantly, 1.32 times more fatal events occurred at home as compared to certificates registered in the five previous years. This suggests that a substantial proportion of the estimated excess mortality during the first wave of COVID-19 resulted from pandemic-induced avoidance of hospitals and under-utilization of healthcare resources for both non-COVID-19 acute conditions [22, 23] and chronic diseases [24, 25]. This observation reflects the reality of overburdened hospitals during the pandemic and aligns with evidence that patients with less urgent needs were sometimes sent back home [26]. Simultaneously, excess all-cause mortality in hospitals was 1.27 times higher than in the five previous years in Colorado. A possible explanation could be that higher occupancy rates led to higher in-hospital mortality rates (e.g., among critically ill [27] and ER patients [28]). Furthermore, excess deaths strongly affected Colorado nursing homes. In assisted living facilities and skilled nursing homes alone, 149 outbreaks were recorded across 19 counties between March 5th and September 10th of 2020, yielding 499 ascertained and probable infections, as well as 172 deaths among residents and staff members. Of note, 57% of nursing home outbreaks were concentrated in three counties, all urban: Denver, Arapahoe, and Jefferson [9]. Conversely, Colorado hospices did not experience excess mortality. During the time period considered, only one outbreak was recorded in such a facility (Alamosa County, July 2020 [9]). This result can be interpreted in light of a decreasing trend in the number of deaths occurring in hospices observed over the last five years, ranging from 42.1 to 32.7 deaths per 100k between 2015 and 2019 (i.e., a reduction by 22.3%).

This study leverages an intersectional approach [29, 30] to measure disparities in both COVID-19 mortality and excess all-cause mortality. Thus, we stratified our analysis by place and sociodemographics – as reflected across age, sex, race, and ethnicity. Such multi-factorial analyses [31, 32] can make a contribution towards health equity by proposing explicit criteria [33], including excess all-cause mortality, to identify population subgroups most vulnerable to the downstream consequences of the COVID-19 pandemic. The strength of excess mortality estimates lies in capturing the pandemic’s impact across causes of death and at different scales. Such a study was made possible through a close collaboration with the CDPHE Office of Vital Records, allowing us to gain access to individual-level death certificates used by the CDC to ascertain whether SARS-CoV-2 infection was the underlying or contributing cause of death. This data-sharing partnership was crucial for our ability to analyze deaths at the county-level and for each population subgroup. We quantified variation in excess mortality across geographies and demographics, and further proposed a preliminary attribution of excess deaths indirectly related to SARS-CoV-2 infection across acute and chronic conditions. The analysis of sub-state level data pinpoints local differences that are often overlooked at an aggregated level [34]. It also provides a quantitative foundation for more targeted policy responses, including implications for directing state and county health budgets towards the most impacted population subgroups.

Further applying such an intersectional approach, we stratified COVID-19 and excess deaths by race, ethnicity, and sex to examine the disparate impacts of COVID-19 in Colorado. Early in the pandemic, the CDPHE published a press release [35] acknowledging the disparities in SARS-CoV-2 infection and mortality, especially with respect to race and ethnicity [36]. Our study confirmed their preliminary observations: Black and Hispanic White males – representing 32% of the population [37] – were most afflicted, with 232 and 186 estimated excess deaths per 100k respectively, from March to September 2020. Our results corroborate previous CDC reports at the national level as well, which demonstrated that COVID-19 has been more deadly among Black and Hispanic population subgroups [38, 39], while providing more detailed estimates by race, ethnicity, and sex. Additionally, we showed that all-cause mortality disproportionately affected younger adults, as measured by the ratio of deaths in 2020 to the average of the five previous years. In other words, although the level of excess deaths in Colorado was larger among older populations in absolute value, as observed across the country [40, 41] during the first COVID-19 wave, working age adults (35-44) experienced the highest all-cause mortality ratio (1.4 times more deaths in 2020, as compared to the five previous years). For comparison, all-cause mortality among older populations (aged 75+) was 1.2 times higher than in 2015 to 2019.

Our study also quantified geographic differences in COVID-19 and excess all-cause mortality in Colorado from March to September 2020, in aggregate and over time. Place-based factors have long been implicated as a root cause of socioeconomic disparities and adverse health outcomes [42, 43, 44, 45]. Three counties were more affected than others in terms of excess all-cause mortality: Morgan, Chaffee, and Dolores. In Morgan County, three large outbreaks in nursing homes and food processing factories may have contributed to the excess death toll during the first COVID-19 wave [46]. Similarly, in Chaffee County, the presence of non-hospital health facilities and meatpacking plants accelerated the local spread of SARS-CoV-2 infection [47]. Finally, in Dolores County, one of the most economically underprivileged in the state (12.8% living under the poverty line [48]), multiple factors may have played a role in excess all-cause mortality, including a large aging population (25% aged 65+) and an under-resourced healthcare infrastructure [49]. Both industry- and population-based factors could explain variation in excess mortality outcomes across counties, e.g., some counties have a higher prevalence of industries with increased risk of exposure, such as meatpacking or oil and gas drilling platforms, and others have a greater concentration of vulnerable populations.

In sum, this study offers a framework to analyze the impact of the COVID-19 pandemic on mortality and explores excess mortality attributable to specific causes of death in Colorado. Nevertheless, four limitations deserve mention. First, despite the increasing use of high-resolution data to uncover local patterns in health outcomes and inform public health efforts [50, 51, 52, 53], the estimation of noisy excess all-cause mortality rates might err in counties with small populations (< 1000 residents). Yet, aggregating observations at the CBSA level generally smooths these potential artifacts. Second, future methodological improvements could incorporate temporal trends in all-cause and cause-specific mortality as well as apply a hierarchical Poisson regression modeling framework [54] to analyze death certificates. This approach has been found effective in capturing a high variability in count datasets. In this study, we favored an empirical approach relying on absolute and relative mortality differences to facilitate interpretability. Third, with respect to the accurate determination of the underlying cause of death, a lack of available testing may have hindered the detection of fatal events directly due to SARS-CoV-2 infection early in the pandemic. This underdetection was especially true in nursing homes and other non-hospital health institutions [55, 56, 57, 58], thus affecting counties and CBSAs with more such facilities that were first hit by outbreaks. Adding to the uneven geographic accessibility to testing [59], changes in testing capacity and practice might in turn influence the estimation of excess all-cause mortality attributable to SARS-CoV-2 infection over time, *directly* or *indirectly*. Nonetheless, these effects can be mitigated by adopting a cross-sectional approach and analyzing the impact of each COVID-19 wave on all-cause mortality separately. Fourth, some of our results might be attributable to the specific context in Colorado. Geographical variation in the proportion of excess deaths directly due to SARS-CoV-2 infection, as opposed to other acute and chronic fatal events, remains to be determined beyond the case of that state. As reflected by prior work on social and epidemiological vulnerability [60, 61], there is indeed a high variation in healthcare system capacity, strength, accessibility, and preparedness across US counties. Yet, our proposed analysis framework is generalizable and could be applied in other states to evaluate statistical variability in excess all-cause mortality at the local level.

In the future, we aim to estimate the effect of local policies and health system capacity on all-cause mortality during the pandemic, and to assess the association between excess mortality and underlying county vulnerability to medical care deficiencies [62, 63, 64]. Furthermore, we seek to better understand the mix of significant contributing factors that led to deaths from SARS-CoV-2 as the direct cause, and plan to investigate the distribution of underlying causes of death among patients with COVID-19 listed as secondary on their certificates. More broadly, downstream repercussions of NPIs such as stay-at-home orders on routine care, chronic diseases, and socioeconomic factors – tied to mental health [65, 66] and well-being [67, 68, 69] – are evident and deserve to be carefully studied to better inform potential courses of action.

## 6. Conclusion

To our knowledge, this study provides the first in-depth analysis of both regional and racial-ethnic disparities in the direct and indirect impacts of the COVID-19 pandemic. We assessed geographic and sociodemographic differences not only in COVID-19 mortality but also in excess all-cause mortality during the first six months of the pandemic in Colorado, as compared to the same period in 2015 to 2019. Such an intersectional approach – capturing the combined effects of sex, race, ethnicity, and place – can serve as the quantitative foundation for more tailored community-based responses to the pandemic at the local level (e.g., by reallocating county health budgets to the most afflicted population subgroups). Our investigation of individual-level death certificate data, obtained thanks to a close collaboration with the CDPHE, lends insight into both the *direct* and *indirect* impacts of the novel SARS-CoV-2 infection in the most vulnerable urban, rural, and frontier counties. More work is needed to determine the contribution of ongoing, structural factors to differential levels of excess mortality (e.g., systemic disparities in economic and health infrastructure), as opposed to situational, pandemic-related factors (e.g., difficult-to-contain outbreaks in congregate settings leading to community transmission). This analysis is a first step toward that end, and in the future, inference studies building upon our proposed framework may elucidate the causal attribution of excess mortality at various geographical resolutions.

## Supporting information

All Supplemental Material

## Data Availability

All analyses were run using Python 3.7.1. All scripts used for analyses, as well as aggregated data, are located on the following Github repository: https://github.com/jaychandra3/excess_mortality_CO.

https://github.com/jaychandra3/excess_mortality_CO

## 8. Acknowledgments

We thank Anna Miller and Alexa Shyama for assistance with data requests and background research. Additionally, we thank Amy Cheung for her thoughtful writing suggestions.

## Notes

### Competing Interest Statement

The authors have declared no competing interest.

### Funding Statement

No external funding was recieved.

### Author Declarations

All data was obtained in accordance with CDPHE's policy for data sharing.

