## Supplementary material for "Evaluation of County-Level Heterogeneity in Excess Mortality in Colorado from March to September 2020": All Supplemental Material

### Supplementary Materials

#### 1. Comparison between COVID-19 data from the JHU CSSE and the CDPHE

To contrast estimates of deaths directly due to SARS-CoV-2 infection as certified by the Colorado Department of Public Health and Environment (CDPHE) with open access datasets available to the general public, we used the COVID-19 death time series from the data repository operated by the Johns Hopkins University Center for Systems Science and Engineering (JHU CSSE) [1]—a monitoring project funded through a RAPID NSF grant [2]. Notably, the JHU dataset counts every death record mapping to a positive COVID-19 test as a COVID-19 death, whereas ascertained death certificates shared by CDPHE and verified by the CDC distinguishes COVID-19 as the underlying vs. contributing but secondary cause of death. These two distinct definitions of COVID-19 death may lead to different estimates of COVID-19 mortality rates. Because the JHU dataset counts every death linked to a positive COVID-19 test as a COVID-19 death, using this definition may result in an overestimation of mortality directly due to the novel SARS-CoV-2 infection. In contrast, using the CDPHE dataset would provide a more reliable assessment of mortality directly due to the novel SARS-CoV-2 infection, since the distinction between underlying and contributing causes of death would be made by the medical examiner completing the official death certificate.

We conducted a longitudinal comparison of the two data sources at both the state (Section 4.1) and county level (Supp Materials 2, Supp Fig. 1, Supp Fig. 2). Note that smaller discrepancies at the county level (from 1 to 83 COVID-19 deaths) sum to a more noticeable distinction (345 COVID-19 deaths) when aggregated to the state level.

#### 2. Longitudinal analysis by county type

A longitudinal analysis of excess all-cause mortality at the county level may lead to data artefacts due to a low population size in certain counties of Colorado. Stratifying by county type will overcome the issues of counties with small population sizes, while maintaining a granular resolution at the county level. Such an analysis would also elucidate the heterogeneous impact of the COVID-19 pandemic across geographies and demographics. The American Communities Project based at The George Washington University's School of Media & Public Affairs classifies United States counties into 15 county types [3]. In Colorado, eight different county types are represented, including Exurbs, Greying America, Urban Suburbs, College Towns, Big Cities, Military Posts, Rural Middle America, Hispanic Centers, and Working-Class Country. Of note, exurbs, Urban Suburbs and Big Cities make up a large portion (62%) of the population in Colorado. These include Denver, Arapahoe, and Broomfield. However, most of the counties in Colorado (58%) are classified as Greying America because of a large proportion of seniors in their population.

Population-normalized excess all-cause mortality was the highest in Urban Suburbs and Big Cities. Though high in Rural Middle America and Hispanic Centers as well, the magnitude of excess deaths was less precisely estimated in such places because of a lower or more variable population size. College towns such as Boulder and Larimer have presented lower magnitudes of excess deaths and COVID-19 mortality, despite having large populations. This might be due to a large portion of the student population leaving campus to go back home. The military post in El Paso county has shown an increase in the cumulative difference between excess deaths and deaths directly due to SARS-CoV-2 infection from

April to September, coinciding with outbreaks reported in military environments. One would expect Greying America to have elevated excess all-cause mortality, given large nursing home outbreaks experienced in Colorado. However, Greying America counties did not exhibit significantly different excess mortality estimates from other county types (Supp Fig. 1).

#### 3. Formula for excess mortality

With a constant population (as of 2017):

$$ED_j = \frac{1}{P_{2017,j}}\left(D_{2020,j} - \frac{1}{n}\sum_{i=2015}^{2019} D_{i,j}\right)$$

With a time-varying population:

$$ED_j = \frac{D_{2020,j}}{P_{2020,j}} - \frac{1}{n}\sum_{i=2015}^{2019}\frac{D_{i,j}}{P_{i,j}}$$

$ED_j$: population-normalized excess deaths in geographical unit j
$D_{i,j}$: number of deaths in year I in geographical unit j
$P_{i,j}$: population in year I in geographical unit j
n: number of baseline years considered (n=5 in this study)

We needed a baseline number of deaths that would be expected without the pandemic. This was defined by averaging the number of deaths recorded in 2015-2019 during the same time periods. We then subtracted this baseline average from deaths registered in 2020, over the corresponding time period. We chose to use this definition in contrast to more complex approaches due to challenges involved with developing estimates for a number of smaller counties in Colorado.

#### 4. Definition of confidence intervals for excess mortality expressed as a difference or ratio

Excess mortality expressed as a difference (standard CI):

$$95\%\ CI = \bar{x} \pm t\frac{s}{\sqrt{n}}$$

Excess mortality expressed as a ratio (CI obtained using the Delta method):

$$95\%\ CI = \frac{1}{\bar{x}} \pm t\frac{s}{\sqrt{n}\cdot\bar{x}^2}$$

$\bar{x}$: average death count in baseline years (2015-2019)
s: standard deviation of death count in baseline years (2015-2019)
n: number of baseline years considered (n=5 in this study)
t: t-statistic for an alpha-level of 0.05 and 4 degrees of freedom (t=2.776)

#### 5. CBSA-level analyses

Core based statistical areas (CBSA) can provide an intuitive understanding as they reflect the geography where people live, work, and interact, and are thus more representative of human behavior.

CBSAs were classified as metropolitan or micropolitan according to the US Census Bureau based on the number of urbanized areas [4]. The 2010 standards provide that each CBSA must contain at least one urban area of 10,000 or more inhabitants. Each metropolitan statistical area must have at least one urbanized area of 50,000 or more inhabitants, while each micropolitan statistical area must have at least one urban cluster of at least 10,000 but less than 50,000 inhabitants. Analysis at both the county and CBSA level can provide valuable insights for policymakers, as governmental entities exist at both levels. A single CBSA's governing body can span multiple counties, such as CBSAs in Colorado Springs, Glenwood Springs, and Montrose in Colorado.

Analysis at the CBSA level confers several benefits, including a more cohesive geographical area that often better represents an economic region within which most mobility occurs. Additionally, further aggregation at the CBSA level helps smooth out some of the artefacts associated with the small population size in some counties, and alleviates the difficulty of interpreting estimates, especially in rural and frontier areas. These motivated our choice of conducting a sensitivity analysis at the CBSA level (Supp Fig. 2). There are 17 CBSAs in Colorado, including 7 metropolitan and 10 micropolitan areas. The largest CBSA in population size is composed of Denver and 9 surrounding counties (10 areas in total), while 13 of the CBSAs only have one county. CBSA-level deaths were obtained by summing all deaths from counties within the CBSA.

From both the cross-sectional and cumulative perspectives, we observed similar results at the CBSA level. Furthermore, the aggregation of counties into larger geographical areas representing contiguous regions of mobility resulted in less heterogeneous mortality outcomes across areas. In short, all 17 CBSAs recorded more excess deaths from all causes than deaths directly due to SARS-CoV-2 infection in March-September 2020. Fort Morgan, Greeley, and the Denver metropolitan area were the most hit by deaths directly due to SARS-CoV-2 infection. Though early pandemic response in the US focused on large, densely populated metropolitan areas, excess mortality estimates at the CBSA level revealed that micropolitan areas such as Craig, Sterling, and Fort Morgan, were most affected by both *direct* and *indirect* COVID-19-related mortality.

From March 5th to September 10th, all CBSAs presented a positive number of excess deaths per 100k person, leaving cluster C being empty, 14 of the 17 CBSAs (83% of CBSAs) presented greater excess deaths per 100k than COVID-19 deaths per 100k (cluster A), and 3 of the 17 CBSAs (17% of CBSAs) presented greater COVID-19 per 100k than excess deaths per 100k (cluster B). The areas primarily affected by deaths directly due to SARS-CoV-2 infection, as ascertained by the CDPHE, were Fort Morgan, Greeley, and Denver-Aurora-Lakewood, i.e., one micropolitan and two metropolitan areas, with 145, 45, and 38 COVID-19 per 100k, respectively. In turn, Pueblo (96 deaths per 100k), Denver-Aurora-Lakewood (86 deaths per 100k), and Grand Junction (80 deaths per 100k) among metropolitan areas, as well as Craig (163 deaths per 100k), Sterling (128 deaths per 100k), and Fort Morgan (107 deaths per 100k) among micropolitan areas were characterized by the highest rates of excess mortality from all causes (Supp Fig. 3).

#### 6. Sensitivity analysis accounting for time-varying population sizes

Excess deaths were calculated for each geographical level (i.e., county, CBSA, and state) in Colorado and normalized by population size. Two different methods were used to account for the size of the population per county. The second approach implemented time-varying county and CBSA population sizes. In order to account for the increasing population in certain areas of Colorado, and in the natural increase in deaths that follows from older populations of increasing sizes in certain counties elected by retirees, we conducted a sensitivity analysis. We divided yearly death counts by the size of the population as estimated for the year prior. Similar to the first approach, population-normalized death counts from years 2015-2019 were averaged and subtracted from 2020 population-normalized deaths, yielding population-normalized excess deaths.

#### 7. Overview of Colorado's population

In 2017, the median age of Colorado residents was 36.8. The projected median age grew from 36.7 in 2015 to 37.3 in 2020. 50% of Colorado's population is Male and 22% of Colorado's population is Hispanic. 89% of the population is White, 5% of the population is Black/African American, 4% of the population is Asian/Pacific Islander, and 2% of the population is American Indian/Alaskan Native. Notably, the median age of the White population is 37 years old, while it is 30 among American Indians and Black/African Americans and 33 among Asian/Pacific Islanders. Additionally, the median age of the Hispanic population in Colorado is 28 years old vs. 39 among Non-Hispanics (Supp Table 1).

#### 8. Timeliness of death certificates

Timeliness of death registration and cause of death certification can vary across states, as detailed in the NCHS report [5]. In Colorado, a three-month lag is considered to be standard for interpreting such data: for example, data through the second quarter of 2020 (January-June) would be virtually complete by the end of September 2020. This would imply that certain death certificates corresponding to fatal events which occurred between July and September might not have been registered yet, depending on the cause of death, such as those from overdose, suicide, and homicide due to ongoing tests [5]. These are often registered with a 'pending' (as opposed to a 'complete' certificate reflecting a certified cause of death) cause of death statement from the medical certifier (i.e., physician, coroner, or medical examiner): the certificate would still be sent to the CDC because of the requirement for death notification within 10 days. It is unusual to provide some cause of death information and add more later. Rather, most death certificates remain 'pending' until the certifier has completed review or investigation.

#### 10. Supplementary Figures

**Supplementary Figure 1. Weekly excess deaths and COVID-19 deaths among residents from February 1st to September 6th, grouped by county type.**

**(a)** Weekly deaths recorded since February 1st, for each of the three time series. **(b)** Cumulative deaths recorded since February 1st, for each of the three time series. For both **(a)** and **(b)**, the blue line represents the estimated excess deaths among county type residents, normalized to the county type's population size projection in 2017. The orange line represents COVID-19 deaths in the county type as reported by the JHU COVID-19 data repository, normalized similarly. The green line represents deaths directly due to SARS-CoV-2 infection in the county type as reported by the Colorado CDPHE, normalized similarly.

**(a)**

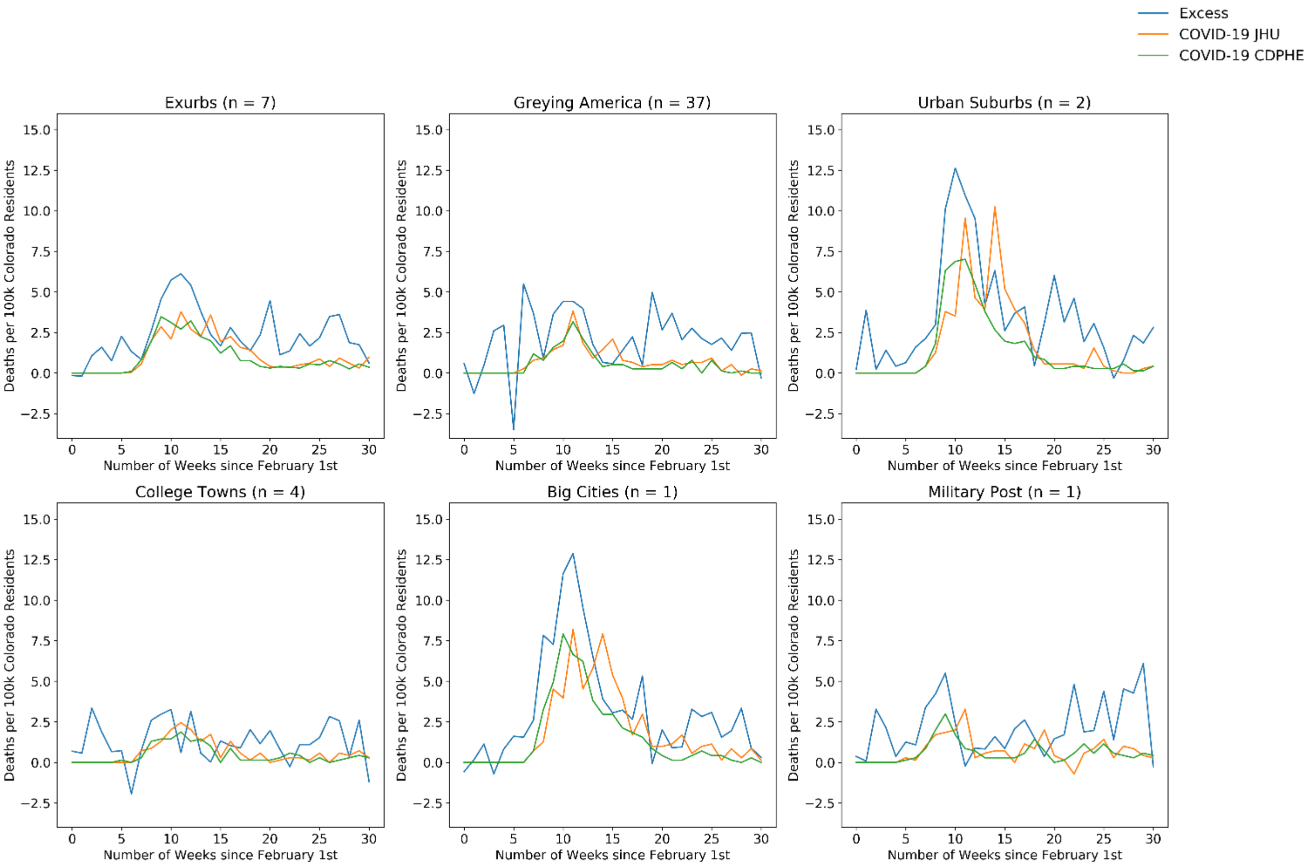

**(b)**

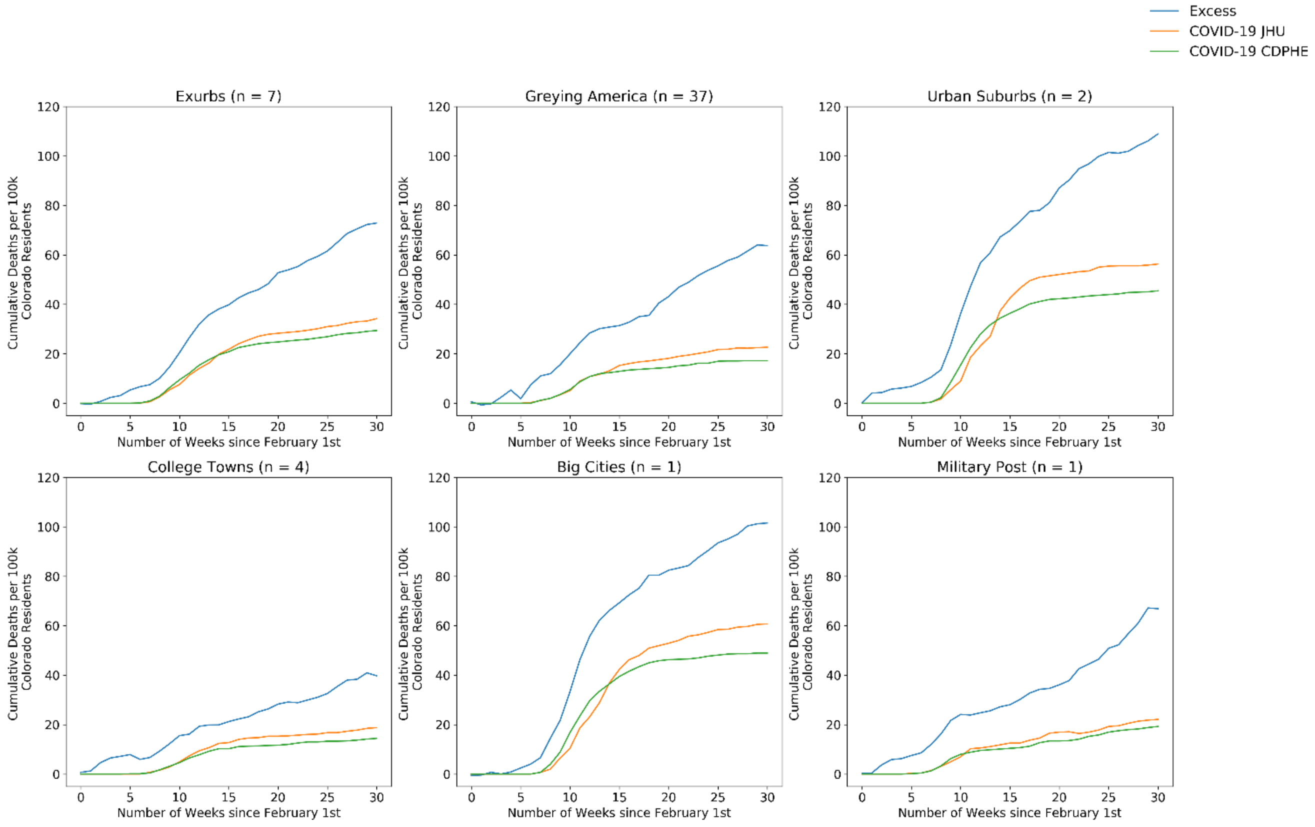


**Supplementary Figure 2. Weekly excess deaths and COVID-19 deaths among residents from February 1st to September 6th, grouped by CBSA.**

**(a)** Weekly deaths recorded since February 1st, for each of the three time series. **(b)** Cumulative deaths recorded since February 1st, for each of the three time series. For both **(a)** and **(b)**, the blue line represents the estimated excess deaths among the CBSA's residents, normalized to the CBSA's population size projection in 2017. The orange line represents COVID-19 deaths in the CBSA as reported by the JHU COVID-19 data repository, normalized similarly. The green line represents the deaths in the CBSA directly due to SARS-CoV-2 infection as reported by the Colorado CDPHE, normalized similarly.

**(a)**

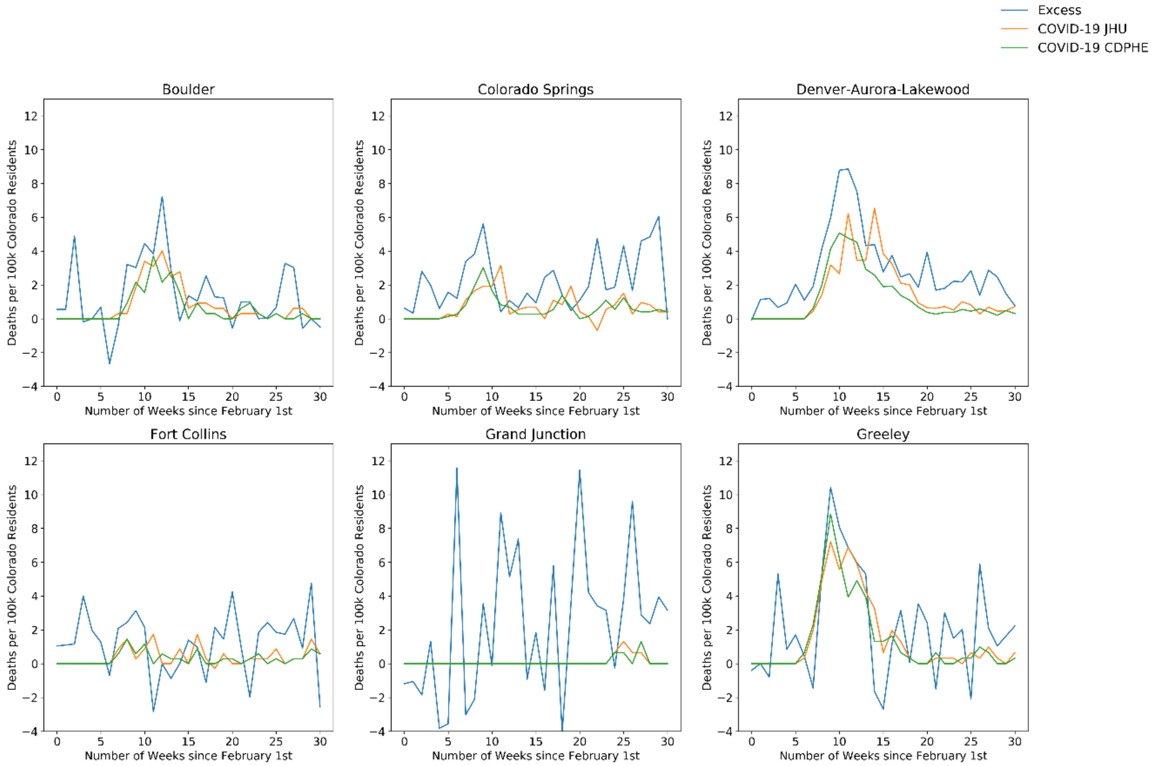
Excess
COVID-19 JHU
COVID-19 CDPHE
Boulder
Colorado Springs
Denver-Aurora-Lakewood
Fort Collins
Grand Junction
Greeley
Deaths per 100k Colorado Residents
Number of Weeks since February 1st

**(b)**

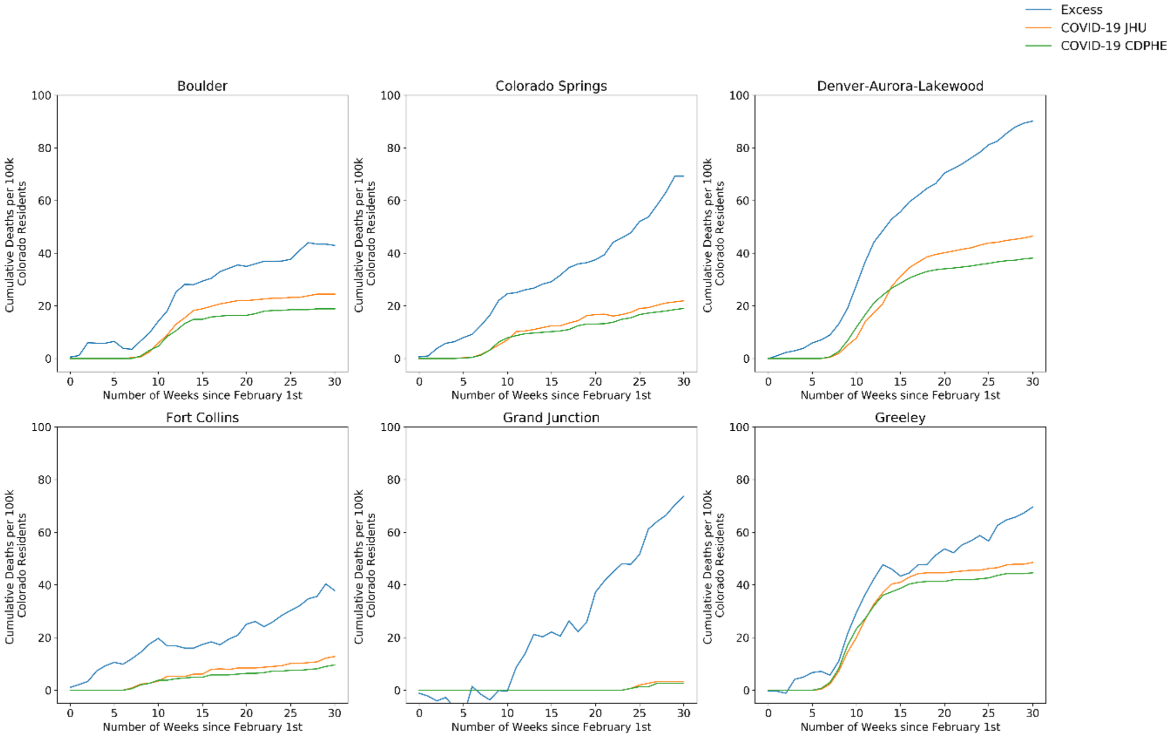
Excess
COVID-19 JHU
COVID-19 CDPHE
Boulder
Colorado Springs
Denver-Aurora-Lakewood
Fort Collins
Grand Junction
Greeley
Cumulative Deaths per 100k Colorado Residents
Number of Weeks since February 1st

**Supplementary Figure 3. Overall CBSA-level excess all-cause mortality vs. deaths directly due to SARS-CoV-2 infection among residents.**

Counts correspond to the period starting on the date of the first documented infection in the state (March 5th) and ending on September 10th. Population-normalized estimates were computed using CBSA-level population projections from 2017. The areas of Colorado that had more population-normalized excess deaths from all causes in 2020 than fatal events primarily attributed to COVID-19 by medical certifiers belong to cluster A. Cluster B is the complement of cluster A. It is composed of areas where the COVID-19 death rate exceeded the value of population-normalized excess all-cause mortality rate. CBSAs belonging to cluster C were characterized by negative excess deaths in 2020 (none of the CBSAs fitted into that category, leaving this cluster empty). The points were sized in proportion to the population of each county and were colored based on the CBSA type (i.e., metropolitan in blue and micropolitan in green).

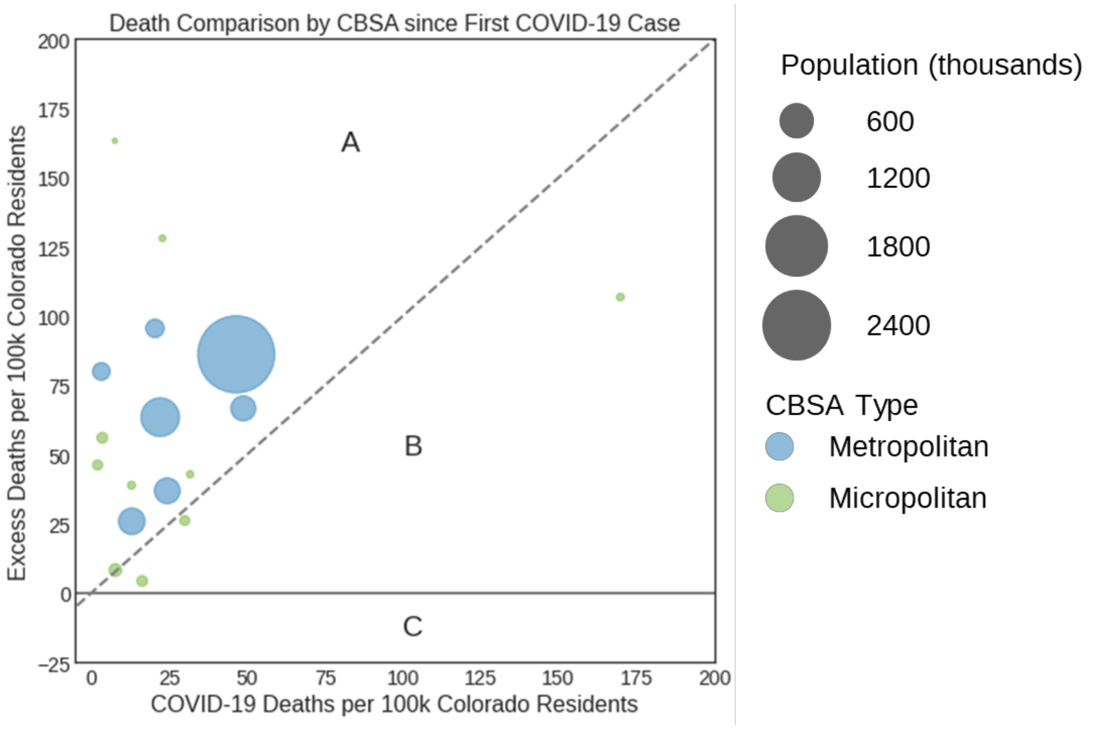


**Supplementary Figure 4. Monthly county-level excess all-cause mortality vs. deaths directly due to SARS-CoV-2 infection among residents, from March to August 2020 (a to f).**

Population-normalized estimates were computed using county-level population projections from 2017. The areas of Colorado that had more population-normalized excess deaths from all causes in 2020 than fatal events primarily attributed to COVID-19 by medical certifiers belong to cluster A. Cluster B is the complement of cluster A. It is composed of areas where the COVID-19 death rate exceeded the value of population-normalized excess all-cause mortality rate. Counties belonging to cluster C were characterized by negative excess deaths in 2020. The points were sized in proportion to the population of each county and were colored based on the county type (i.e., urban in purple, rural in orange, and frontier in green).

(a)

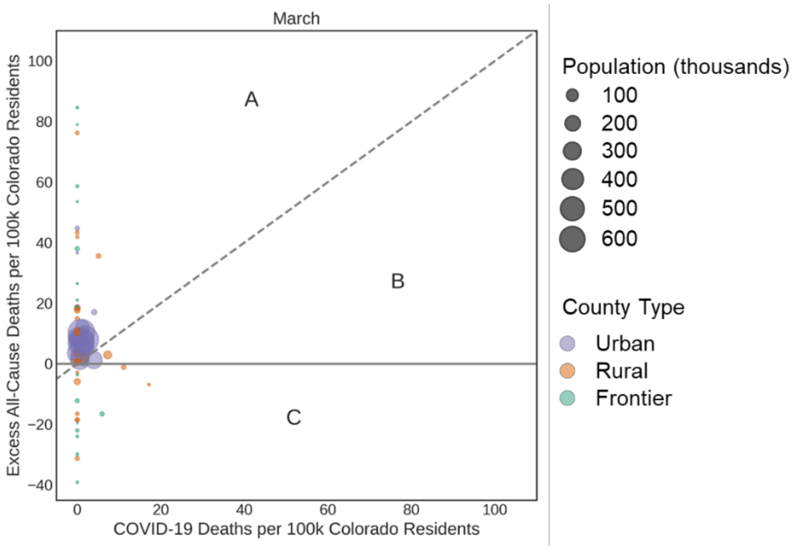

March
Excess All-Cause Deaths per 100k Colorado Residents
COVID-19 Deaths per 100k Colorado Residents
A
B
C
100
80
60
40
20
0
−20
−40
0
20
40
60
80
100
Population (thousands)
100
200
300
400
500
600
County Type
Urban
Rural
Frontier


(b)

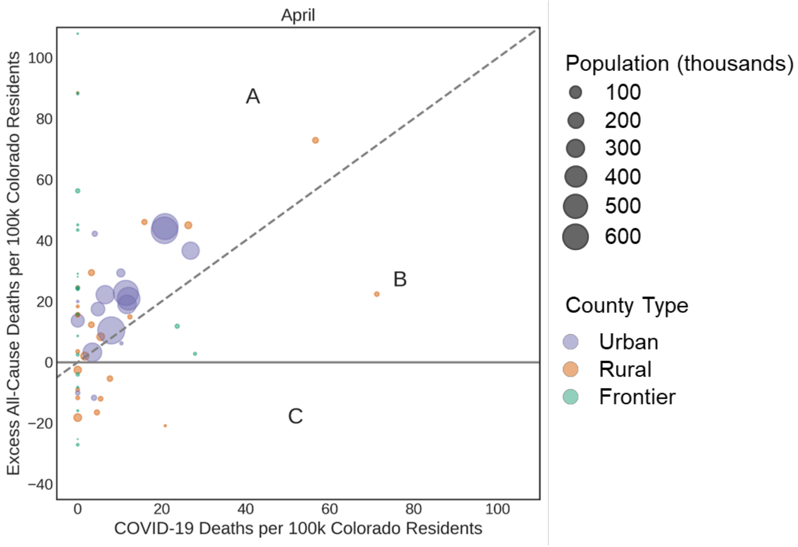

April
Excess All-Cause Deaths per 100k Colorado Residents
COVID-19 Deaths per 100k Colorado Residents
A
B
C
100
80
60
40
20
0
−20
−40
0
20
40
60
80
100
Population (thousands)
100
200
300
400
500
600
County Type
Urban
Rural
Frontier

**(c)**

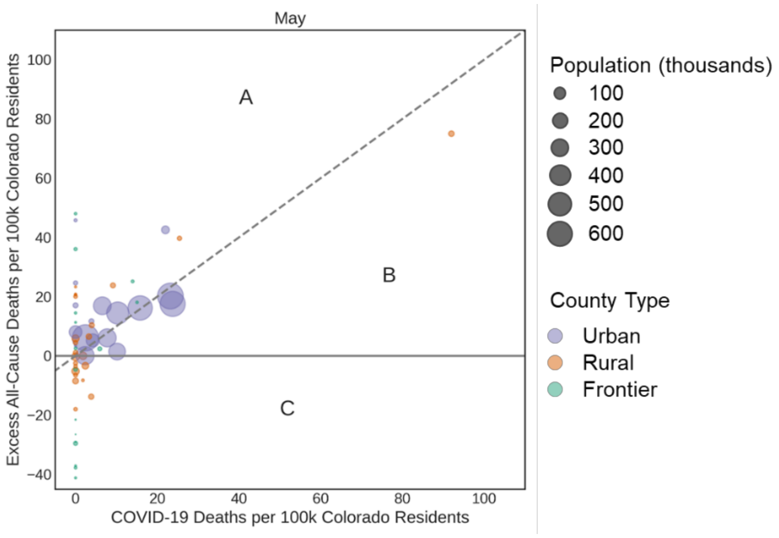
May
A
B
C
Excess All-Cause Deaths per 100k Colorado Residents
COVID-19 Deaths per 100k Colorado Residents
100
80
60
40
20
0
−20
−40
0
20
40
60
80
100
Population (thousands)
100
200
300
400
500
600
County Type
Urban
Rural
Frontier

**(d)**

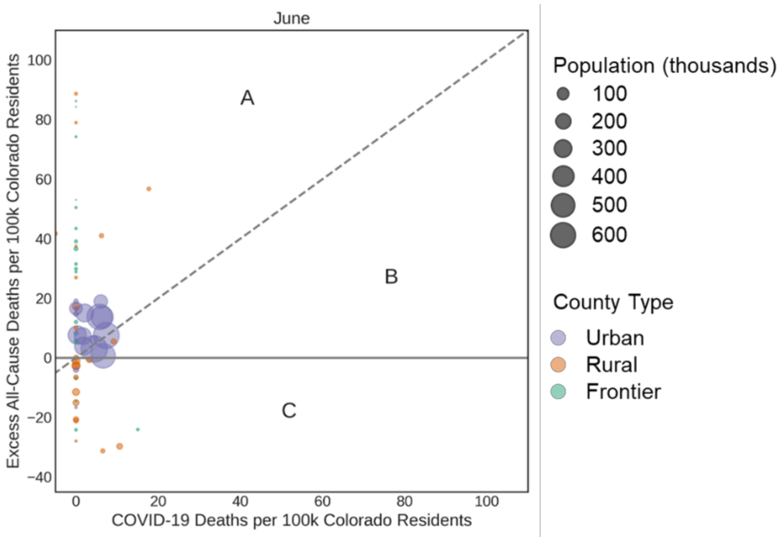
June
A
B
C
Excess All-Cause Deaths per 100k Colorado Residents
COVID-19 Deaths per 100k Colorado Residents
100
80
60
40
20
0
−20
−40
0
20
40
60
80
100
Population (thousands)
100
200
300
400
500
600
County Type
Urban
Rural
Frontier

**(e)**

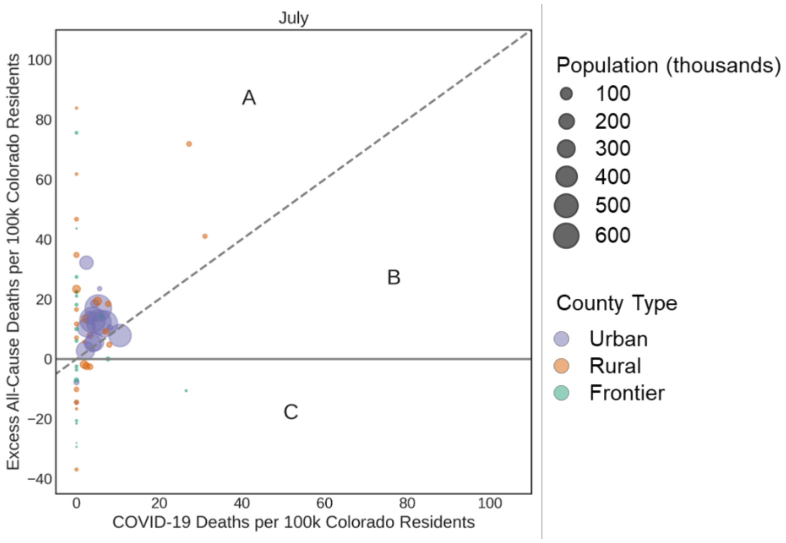
July
A
B
C
Excess All-Cause Deaths per 100k Colorado Residents
COVID-19 Deaths per 100k Colorado Residents
100
80
60
40
20
0
−20
−40
0
20
40
60
80
100
Population (thousands)
100
200
300
400
500
600
County Type
Urban
Rural
Frontier

**(f)**

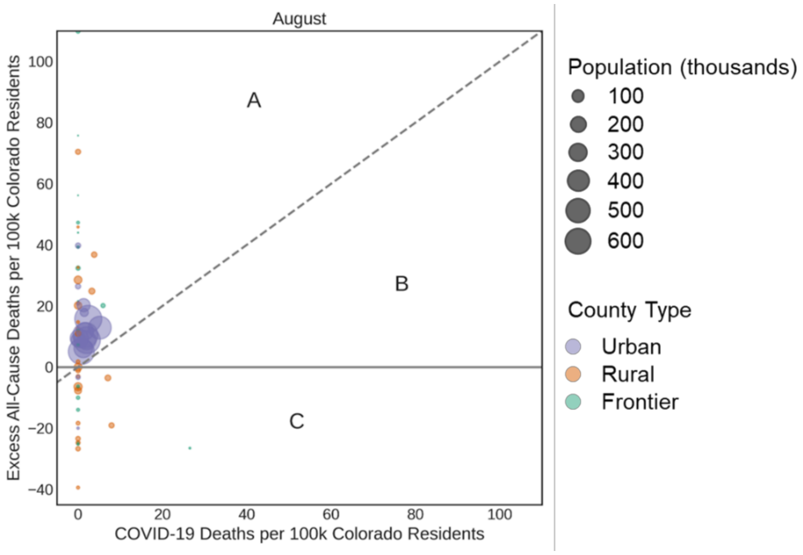


**Supplementary Figure 5. Monthly CBSA-level excess all-cause mortality vs. deaths directly due to SARS-CoV-2 infection among residents, from March to August 2020 (a to f).**

Population-normalized estimates were computed using county-level population projections from 2017. Cluster A includes the areas of Colorado with more excess all-cause deaths than deaths directly due to SARS-CoV-2. Cluster B is the complement of cluster A. It is composed of areas where the COVID-19 death rate exceeded the value of population-normalized excess all-cause mortality rate. In contrast with clusters A and B, cluster C comprises CBSAs characterized by negative excess deaths in 2020. The points were sized in proportion to the population of each CBSA and were colored based on the CBSA type (i.e., metropolitan in blue and micropolitan in green).

**(a)**

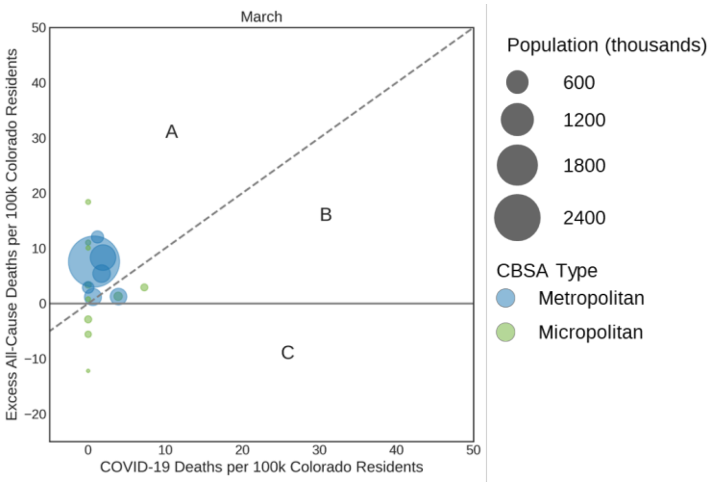

**(b)**

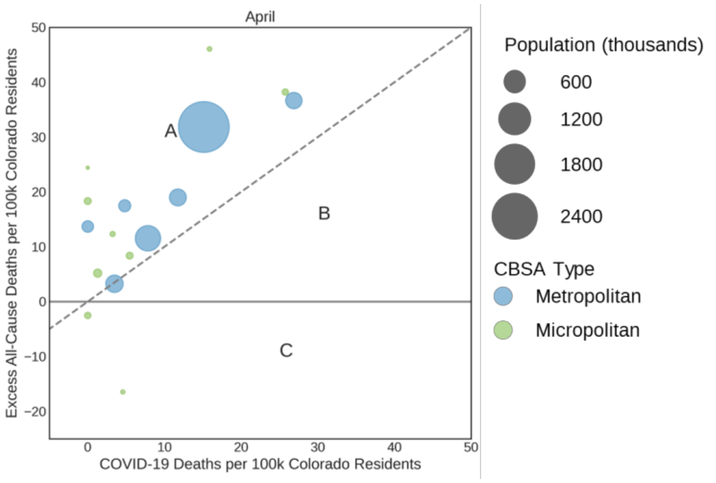
April
Excess All-Cause Deaths per 100k Colorado Residents
COVID-19 Deaths per 100k Colorado Residents
50
40
30
20
10
0
−10
−20
0
10
20
30
40
50
A
B
C
Population (thousands)
600
1200
1800
2400
CBSA Type
Metropolitan
Micropolitan

**(c)**

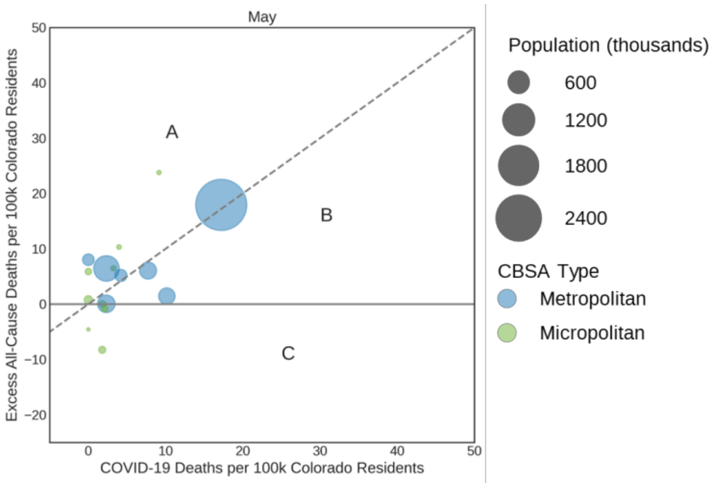
May
Excess All-Cause Deaths per 100k Colorado Residents
COVID-19 Deaths per 100k Colorado Residents
50
40
30
20
10
0
−10
−20
0
10
20
30
40
50
A
B
C
Population (thousands)
600
1200
1800
2400
CBSA Type
Metropolitan
Micropolitan

**(d)**

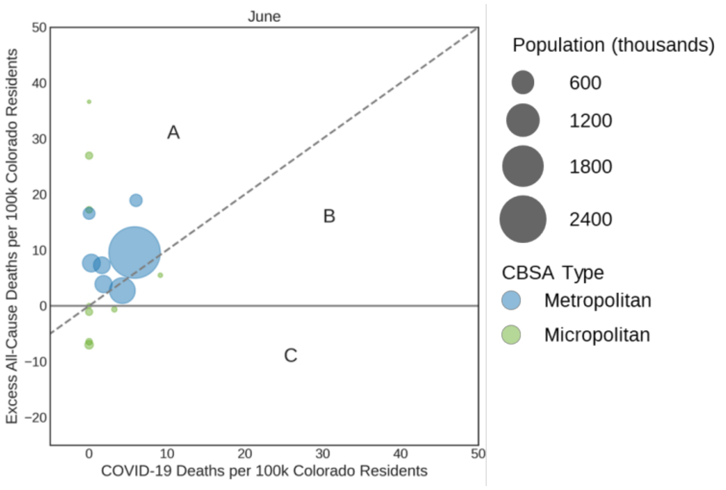
June
Excess All-Cause Deaths per 100k Colorado Residents
COVID-19 Deaths per 100k Colorado Residents
50
40
30
20
10
0
−10
−20
0
10
20
30
40
50
A
B
C
Population (thousands)
600
1200
1800
2400
CBSA Type
Metropolitan
Micropolitan

**(e)**

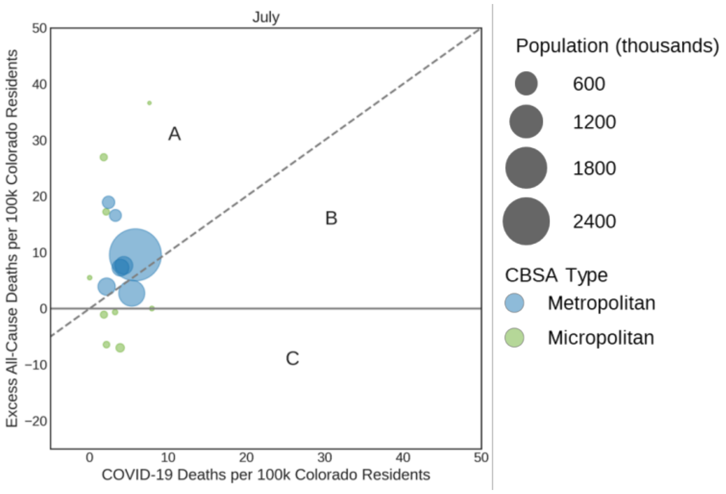


**(f)**

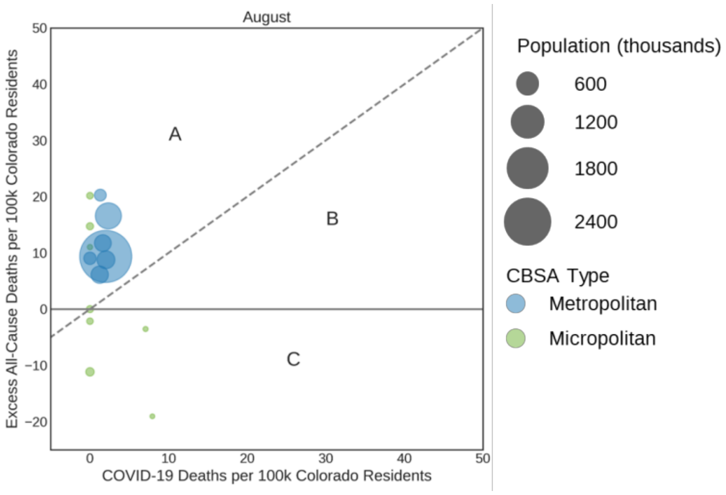


**Supplementary Figure 6. Overall county-level excess all-cause mortality vs. deaths directly due to SARS-CoV-2 infection among residents, when adjusting for the timing of community transmission.**

Counts correspond to the period starting on **(a)** the date of the first documented infection in the state (March 5th), **(b)** 28 days before March 5th (February 6th), and **(c)** 28 days after March 5th (April 2nd) and all ending on September 10th. Population-normalized estimates were computed using county-level population projections from 2017. Cluster A represents the counties of Colorado that had more population-normalized excess deaths from all causes in 2020 than fatal events primarily attributed to COVID-19 by medical certifiers. Cluster B is composed of counties where the COVID-19 death rate exceeded the value of population-normalized excess all-cause mortality rate. Counties belonging to cluster C were characterized by negative excess deaths in 2020. The points were sized in proportion to the population of each county and were colored based on the county type (i.e., urban in purple, rural in orange, and frontier in green).

**(a)**

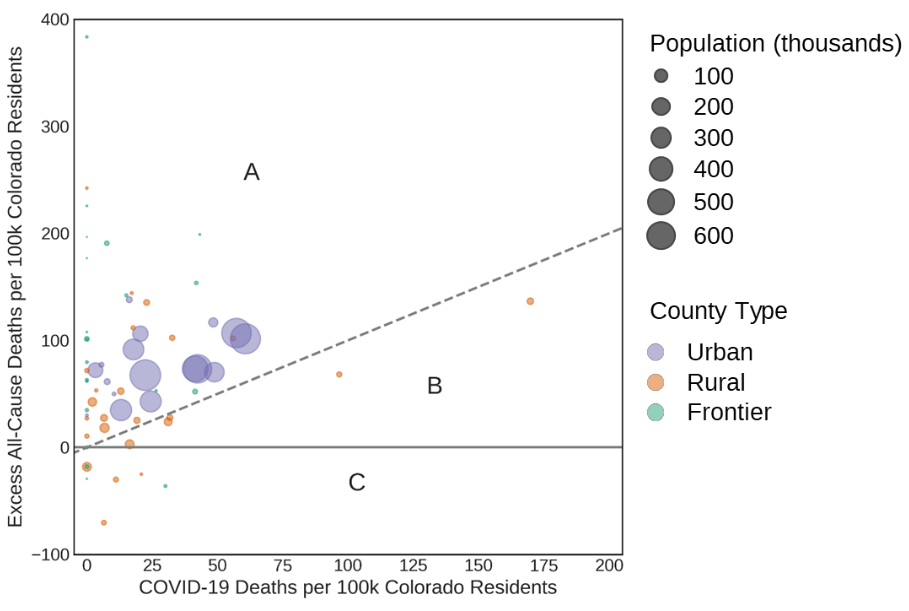

Excess All-Cause Deaths per 100k Colorado Residents
COVID-19 Deaths per 100k Colorado Residents
A
B
C
Population (thousands)
100
200
300
400
500
600
County Type
Urban
Rural
Frontier


**(b)**

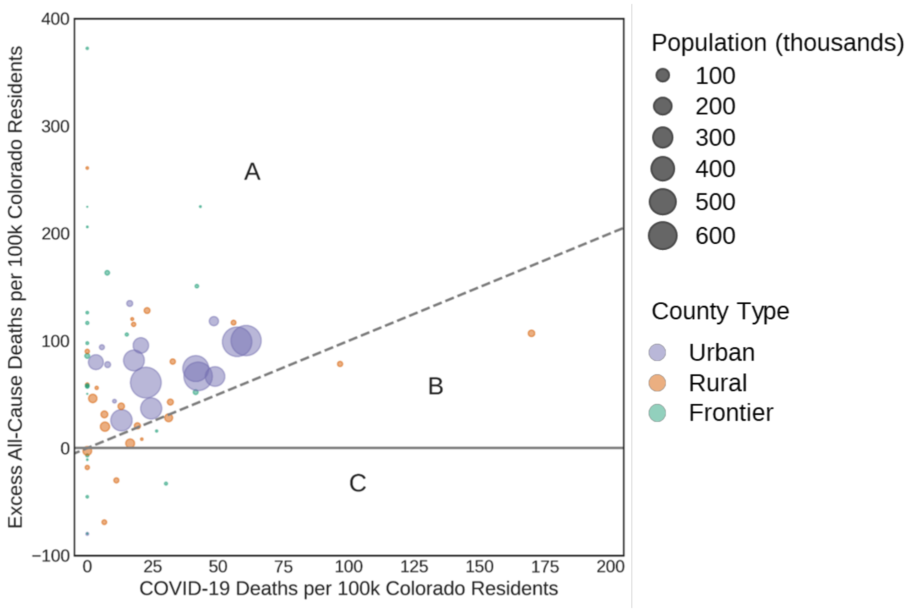

Excess All-Cause Deaths per 100k Colorado Residents
COVID-19 Deaths per 100k Colorado Residents
A
B
C
Population (thousands)
100
200
300
400
500
600
County Type
Urban
Rural
Frontier

**(c)**

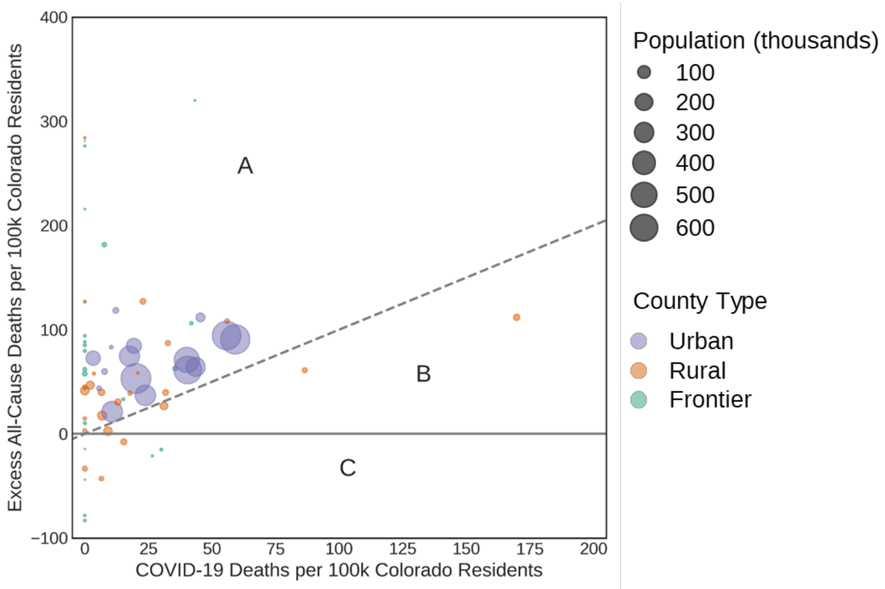


**Supplementary Figure 7. Overall county-level excess all-cause mortality vs. deaths directly due to SARS-CoV-2 infection among residents, with time-varying population normalization.**

Counts correspond to the period starting on **(a)** the date of the first documented infection in the state (March 5th), **(b)** 28 days before March 5th (February 6th), and **(c)** 28 days after March 5th (April 2nd) and all ending on September 10th. Population-normalized estimates were computed by dividing yearly death counts by the size of the population as estimated for the year prior. Cluster A represents the counties of Colorado that had more population-normalized excess deaths from all causes in 2020 than fatal events primarily attributed to COVID-19 by medical certifiers. Cluster B is composed of counties where the COVID-19 death rate exceeded the value of population-normalized excess all-cause mortality rate. Counties belonging to cluster C were characterized by negative excess deaths in 2020. The points were sized in proportion to the population of each county and were colored based on the county type (i.e., urban in purple, rural in orange, and frontier in green).

(a)

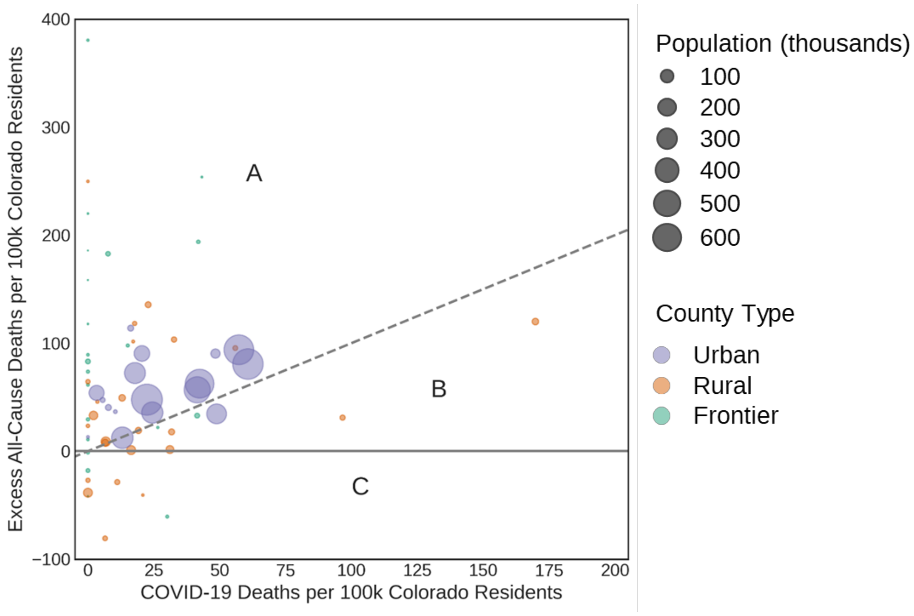

Excess All-Cause Deaths per 100k Colorado Residents
COVID-19 Deaths per 100k Colorado Residents
A
B
C
Population (thousands)
100
200
300
400
500
600
County Type
Urban
Rural
Frontier


(b)

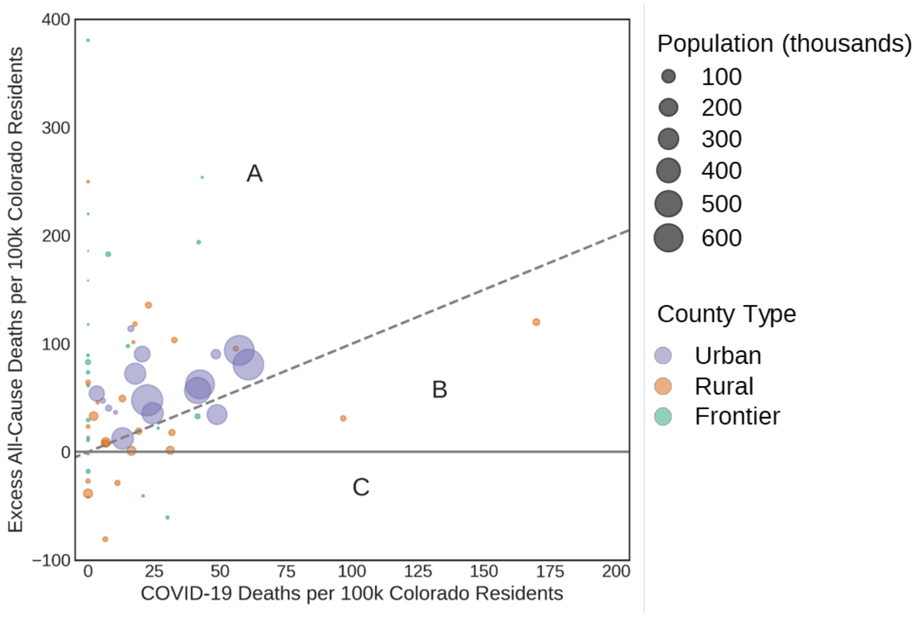

Excess All-Cause Deaths per 100k Colorado Residents
COVID-19 Deaths per 100k Colorado Residents
A
B
C
Population (thousands)
100
200
300
400
500
600
County Type
Urban
Rural
Frontier

**(c)**

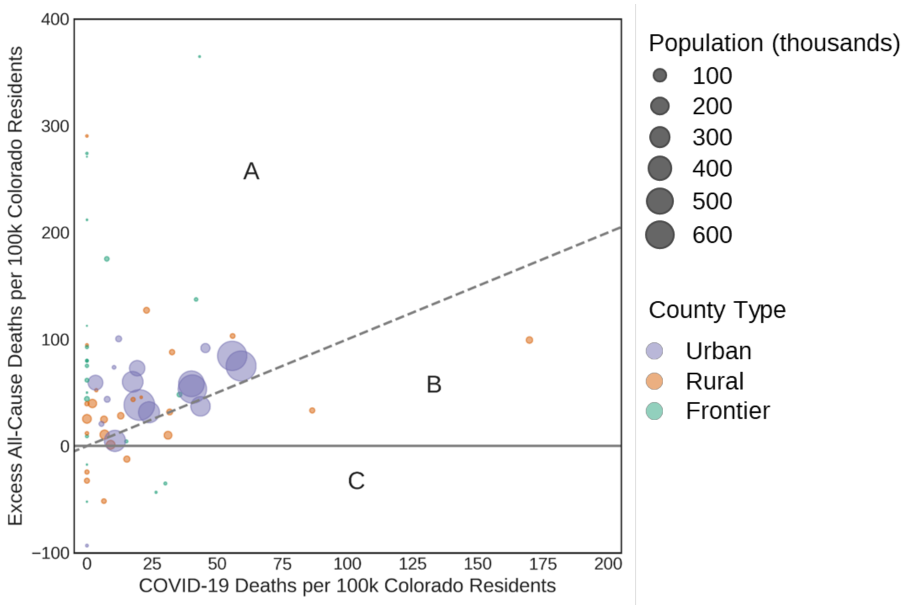


**Supplemental Figure 8. Excess all-cause mortality among residents, stratified by age group.**

Counts correspond to the period starting on the date of the first documented infection in the state (March 5th) and ending on September 10th. Population-normalized estimates for each age group were computed using state-level population projections from 2017. Age groups are presented using ten-year intervals except for the "0" and "1 to 4" age groups which represent newborns/infants and toddlers respectively, and the oldest age group that represents anyone older than 85. **(a)** Age-stratified excess all-cause mortality estimated value and **(b)** age-stratified ratio of deaths in 2020 to the average deaths from 2015 to 2019.

**(a)**

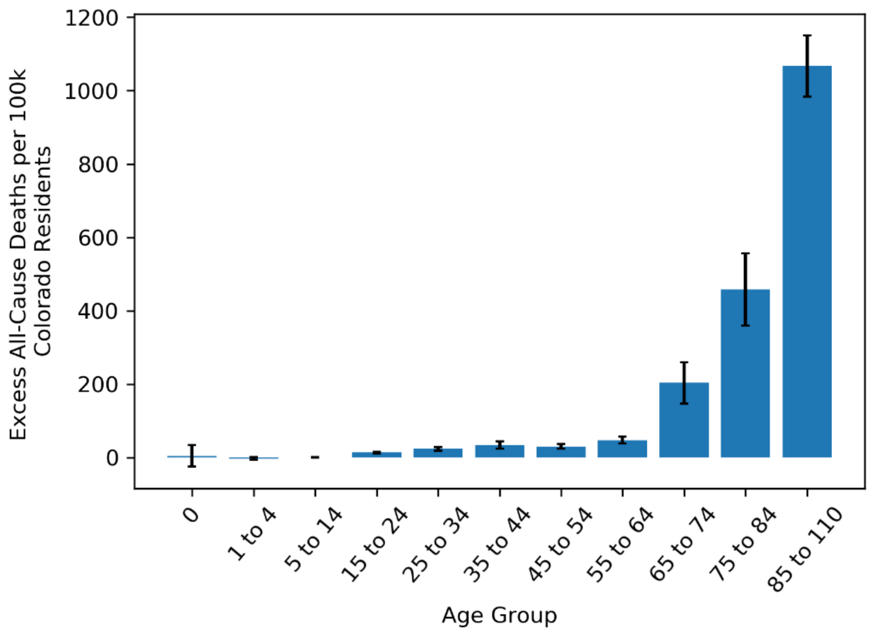

**(b)**

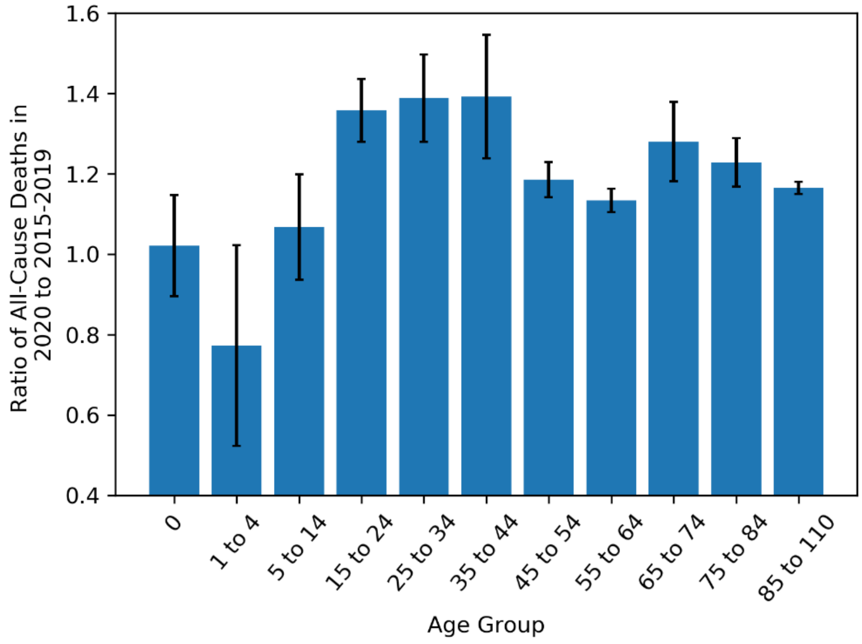


**Supplemental Figure 9. Age-adjusted COVID-19 death rates among residents, stratified by (race, ethnicity, sex) population subgroup.**

Counts correspond to the period starting on the date of the first documented infection in the state (March 5th) and ending on September 10th. Population-normalized estimates were computed using state-level population projections from 2017. The age-adjusted and population-normalized COVID-19 death rates, as reported by the Colorado CDPHE, are shown for each (race, ethnicity, sex) population subgroup in Colorado. Hashed and solid bars represent Hispanic ethnicity, while solid bars represent Non-Hispanic ethnicity. Blue and pink bars represent male and female excess deaths, respectively. Vertically, results are grouped by race (Black/African American, White, Asian/Pacific Islander, and American Indian/Alaska Native).

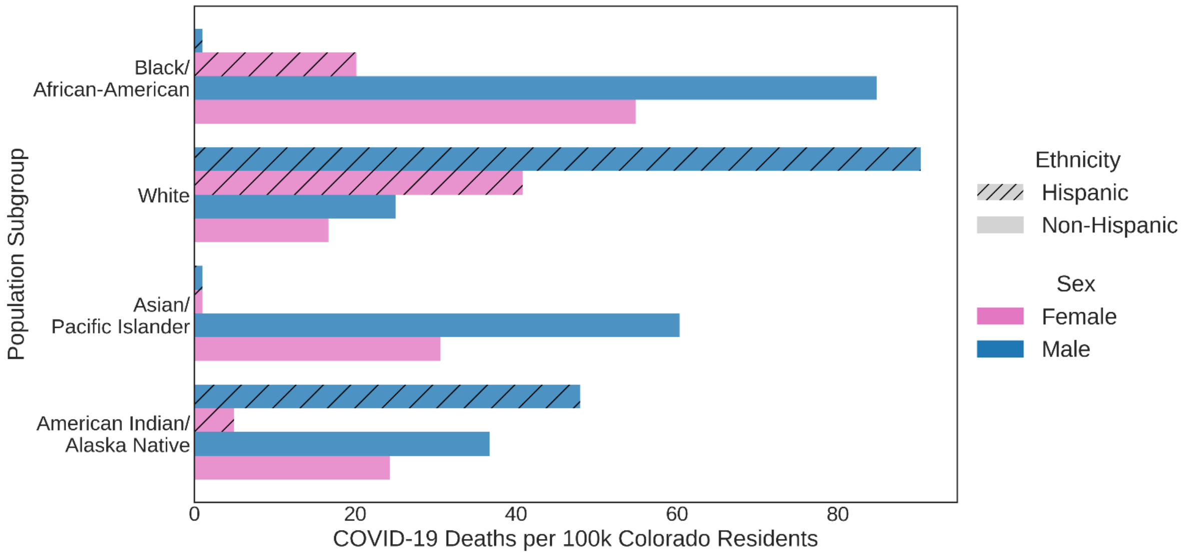

#### Supplementary Tables

**Supplemental Table 1. 2017 Colorado population projections stratified by race, ethnicity, and sex.**

| Race, Ethnicity, Sex | 2017 Population |
|---|---|
| ('American Indian', 'Hispanic Origin', 'M') | 32274 |
| ('American Indian', 'Hispanic Origin', 'F') | 31661 |
| ('American Indian', 'Not of Hispanic Origin', 'M') | 21242 |
| ('American Indian', 'Not of Hispanic Origin', 'F') | 21629 |
| ('Asian/Pacific Islander', 'Hispanic Origin', 'M') | 8506 |
| ('Asian/Pacific Islander', 'Hispanic Origin', 'F') | 8436 |
| ('Asian/Pacific Islander', 'Not of Hispanic Origin', 'M') | 94809 |
| ('Asian/Pacific Islander', 'Not of Hispanic Origin', 'F') | 111503 |
| ('Black', 'Hispanic Origin', 'M') | 18814 |
| ('Black', 'Hispanic Origin', 'F') | 18551 |
| ('Black', 'Not of Hispanic Origin', 'M') | 137570 |
| ('Black', 'Not of Hispanic Origin', 'F') | 121559 |
| ('White', 'Hispanic Origin', 'M') | 554507 |
| ('White', 'Hispanic Origin', 'F') | 539012 |
| ('White', 'Not of Hispanic Origin', 'M') | 1943321 |
| ('White', 'Not of Hispanic Origin', 'F') | 1952314 |

**Supplemental Table 2. ICD-10 codes by cause of death.**

| **Cause of death category** | **List of ICD-10 codes** |
|---|---|
| COVID-19 | U07.1 |
| Motor Vehicle Accident | V02–V04, V09.0, V09.2, V12–V14,V19.0–V19.2, V19.4–V19.6, V20–V79, V80.3–V80.5, V81.0–V81.1, V82.0–V82.1, V83–V86, V87.0–V87.8, V88.0–V88.8, V89.0, V89.2 |
| Total Transport Accidents | V01–V99, Y85 |
| Non-transport Accidents | W00–X59, Y86 |
| Homicide | X85–Y09, Y87.1 |
| Drug Overdose (unintentional and undetermined intent) | X40-X49, Y10-Y14 |
| Intentional Self Harm (suicide) | X60–X84, Y87.0 |
| Pneumonia | J12–J18 |
| Influenza (flu) | J10–J11 |
| Cancer (malignant and non-malignant) | C00-C97, D00-D48 |
| Heart Disease | I00-I09, I11, I13, I20-I51 |
| Cerebrovascular Disease (including stroke) | I60-I69 |
| Chronic Lower Respiratory Disease/COPD (including asthma) | J40-J47 |
| Diabetes Mellitus | E10-E14 |
| Chronic Liver Disease and Cirrhosis | K70, K73-K74 |
| Hypertension (essential/primary hypertension and hypertensive renal disease) | I10, I12 |
| Alzheimer’s Disease | G30 |
| Kidney Disease (nephritis, nephrotic syndrome, and nephrosis) | N00-N07, N17–N19, N25, N27 |
| Work-Related Injuries | Affirmative checkbox on death certificate: “Was the injury work-related?” |
